# Supporting decision-making on allocation of ICU beds and ventilators in pandemics

**DOI:** 10.1101/2020.09.20.20198184

**Authors:** Magnolia Cardona, Claudia C. Dobler, Eyza Koreshe, Daren K Heyland, Rebecca Nguyen, Joan P.Y. Sim, Justin Clark, Alex Psirides

**Affiliations:** Institute for Evidence-Based Healthcare, Bond University Gold Coast, Queensland, Australia; Gold Coast University Hospital, Southport, Queensland, Australia; Evidence-Based Practice Center, Robert D. and Patricia E. Kern Center for the Science of Health Care Delivery, Mayo Clinic, Minnesota, USA; Boden Institute, University of Sydney, NSW, Australia; Department of Critical Care Medicine, Queens University, Kingston, Ontario, Canada; The University of New South Wales, South Western Sydney Clinical School, NSW Australia; Intensive Care Unit, Wellington Regional Hospital, Wellington, New Zealand

**Keywords:** intensive care, ventilator, COVID-19 pandemic, decision-making, healthcare rationing, triage, review

## Abstract

As the world struggles with the COVID-19 pandemic, health service demands have increased to a point where healthcare resources may prove inadequate to meet demand. Guidelines and tools on how to best allocate intensive care beds and ventilators developed during previous epidemics can assist clinicians and policy-makers to make consistent, objective and ethically sounds decisions about resource allocation when healthcare rationing is inevitable. This scoping review of 62 published guidelines, triage protocols, consensus statements and prognostic tools from crisis and non-crisis situations sought to identify a multiplicity of objective factors to inform healthcare rationing of critical care and ventilator care. It also took ethical considerations into account. Prognostic indicators and other decision tools presented here can be combined to create locally-relevant triage algorithms for clinical services and policy makers deciding about allocation of ICU beds and ventilators during a pandemic. Community awareness of the triage protocol is recommended to build trust and alleviate anxiety among the public. This review provides a unique resource and is intended as a discussion starter for clinical services and policy makers to consider formalising an objective triage consensus document that fits the local context.

**Take-home message:** An evidence-based catalogue of objective variables from 62 published resources tested in crisis and non-crisis situations can help clinicians make locally relevant triage decisions on ICU and ventilator allocation in inevitable COVID-19 health rationing.

## Background

The COVID-19 pandemic stretched hospital resources to their limits and beyond in Italy, Spain, England, France, Brazil and the United States [1].Countries and cities that were not affected too badly initially may experience ongoing epidemic waves, and some countries are likely to reach maximum hospital capacity in the near future. A number of countries have faced a shortage of ventilators for COVID-19 patients due to pre-pandemic deficiency. There has been demand for additional care sites and health care to handle a surge in COVID-19 patients. Reported hospitalisation rates are age related, ranging from 0-1% (in people aged 29 years or younger) to 80% (in people aged 80+ years) [2]. Of hospitalised patients, 4.6 to 45.9% have required treatment in the ICU [2-4]. Of all those requiring critical care, 75%, 76% and 88% ended up receiving treatment on a ventilator in the UK, the USA and Italy respectively [5-7]. The length of stay in ICU for COVID-19 patients on ventilators has been longer than in non-crisis periods (median 10 days IQR between 8-14 days in Italy)[7] and median 18 days IQR 9-28 in USA) [8]. Over a quarter (26 to 38%) of those admitted to ICU have died [3,6,7], and overall, people older than 75 years have experienced the highest COVID-19 mortality rates (from 33.5% in UK, to 75.5% in France) [8,9].

ICUs have some capacity to respond to increased demand by surging ICU beds, repurposing hospital spaces, purchasing additional ventilators and hiring and/or training the health workers needed to care for critically ill patients. However, with rapidly increasing and sustained demand such measures might be insufficient, and the need for resources may rapidly exceed capacity. In this situation, healthcare systems need to have evidence-based, equitable, and publicly defensible policies in place on how to ration potentially life-saving treatments [10,11]. Rules to guide allocation of life sustaining treatments will need to incorporate ethical considerations such as social justice, non-prejudice, prevention of preferential treatment of population subgroups, and be transparent to clinicians and the public to prevent moral distress and outrage.

While many health services have ICU admission policies in place for routine care, a recent survey of US services found vast heterogeneity in ventilator triage policies for COVID19. Policies were based (exclusively or in combination) on subjective perceptions of benefits to patients and medical need, ethical considerations, and objective clinical scoring systems [12]. A common gap in these recommendations is the lack of integration of patient values and treatment preferences [13].

We conducted a scoping review of publications that provide prognostic prediction tools or models, and/ or objective triage recommendations which can inform allocation of ICU beds and/or ventilators. The specific objectives were:

1. To identify criteria for ICU admission and ventilator allocation used in epidemic situations as well as during routine care
2. To identify prognostic tools used in patient care during and outside epidemic situations that can potentially enhance confidence in decision-making about resource allocation during the COVID-19 pandemic; and
3. To discuss applicability of these tools for ICU triage in future global emergencies

## Methods

For this scoping review, we searched the databases Medline and Embase on 1^st^ May 2020 for English language articles published since 1^st^ January 2002 (the year in which a SARS outbreak emerged) [14]. Additionally, we manually searched institutional websites of professional intensive care societies, reference lists of systematic reviews, pandemic guidelines from World Health Organization and Centre for Disease Control, and consulted with experts in the field (AP, CD, DH). Details of the full search strategy are shown in Supplement 1, Table S1.1

### Inclusion criteria

We included studies that described patient characteristics and clinical or laboratory parameters/algorithms to facilitate healthcare triage **(I)**. Settings included were hospital wards, emergency departments, and ICUs. To be eligible for inclusion, studies had to report either the accuracy of a prognostic tool (area under the receiver operating characteristics curve [AUROC], or sensitivity/specificity), the validity of a prognostic tool, or significant correlation between a prognostic tool and clinical outcomes (mortality or complication rates) using odds ratios and 95% confidence intervals (CI). Target populations **(P)** were adults or children presenting to hospital during pandemic or non-crisis periods. We included cohort studies, retrospective analyses with large sample sizes (>100 patients), case-control studies, position statements of clinical colleges or professional societies, and consensus documents or guidelines. Selected articles were classified as applicable during an epidemic or applicable to routine care (i.e non-crisis situations).

### Exclusion criteria

We excluded pre-prints; recommendations based on i) subjective clinical judgments only; ii)test result cut-off points that were not validated or at least statistically significantly associated with clinical outcomes; iii) disease-specific scores (e.g. for blood cancers or traumatic brain injury); that were irrelevant to COVID-19; recommendations related to injury in mass casualties; modelling studies; studies with a small sample size (<100); conference abstracts with insufficient data on association/prediction performance; poor performing algorithms; publications in non-Western health systems; and reports with a focus on logistics, ethics of surge capacity.

### Outcomes

Our outcomes of interest **(O)** were performance of algorithms or individual clinical or laboratory factors in predicting patient outcomes, such as in-hospital, 30-day, 90-day, or 1-year mortality. Comparators **(C)** for the validation studies were any other algorithm or laboratory test used by authors, or confirmation of outcomes at discharge or follow-up as specified.

### Data extraction and synthesis

Dyads of reviewers (MC, EK, RN, JS), independently extracted the following study information using a standardized data extraction sheet: author, year of publication, study country, publication type, characteristics of target population disease-specific information), whether pandemic or routine care, type of triage decision (for admission to ICU, discharge from ICU), decision algorithms, and characteristics of risk prediction tools. All discrepancies were discussed within the dyads until consensus was achieved. Algorithm components and variables were presented in tabular form according to source from crisis or non-crisis situation, and whether they were used in intensive care, emergency departments or hospital wards. No meta-analysis was attempted, as this was a scoping review.

## Results

Sixty-two publications relating to 20 countries (eight studies covered multiple countries) met the eligibility criteria: Seven pandemic guidelines/consensus papers/frameworks,[11!,15-20] Nine pandemic triage factors/decision algorithms, [8,21-28] and 46 algorithms, prognostic tools or guidelines for non-crisis care met our inclusion criteria: 19 for routine ICU care [29-47], 21 for routine emergency department care [48-68], and six for general ward care [47,69-73]. The screening process is illustrated in Figure 1, and reasons for exclusion of studies are presented in Supplement 1, Table S1.2.

**<Figure 1. PRISMA diagram illustrating inclusions and exclusions> about here**

The target age groups for the pandemic publications were predominantly adults of all ages admitted to ICU, whereas the routine care algorithms included patients in emergency departments or ICU, and three were exclusively for babies and children [32,34,72]. Supplement 1, Table S1.3 summarises the target populations, study types, and context for pandemic situations, and Table S1.4 shows the corresponding information for routine care. Fourteen of the pandemic-related articles and 8 of the routine care papers included factors that applied to patients with pneumonia. Other conditions to which risk prediction tools/factors were applied included influenza, sepsis, acute injuries due to natural disasters in adults and children, exacerbation of chronic obstructive pulmonary disease, chronic kidney disease and heart failure. Most of the pandemic-related publications suggested criteria for ICU admission, ICU discharge or ventilator allocation, although not all addressed all three questions.

### Domains used for decision-making

A summary of references for recommendations to consider in the decision to escalate care, admit individuals to ICU, and allocate ventilators is presented in Table 1. One domain for decision making included variables and scores to determine patients’ need for higher-level care (column A) to patients who may not yet be in ICU, ventilated or receiving other organ support but may do so later (Table 1, column A). Another domain for decision-making included predictors for patients who stand to benefit from ICU care or mechanical ventilation the most and should be prioritised (Table 1, column B).

**Table 1.**
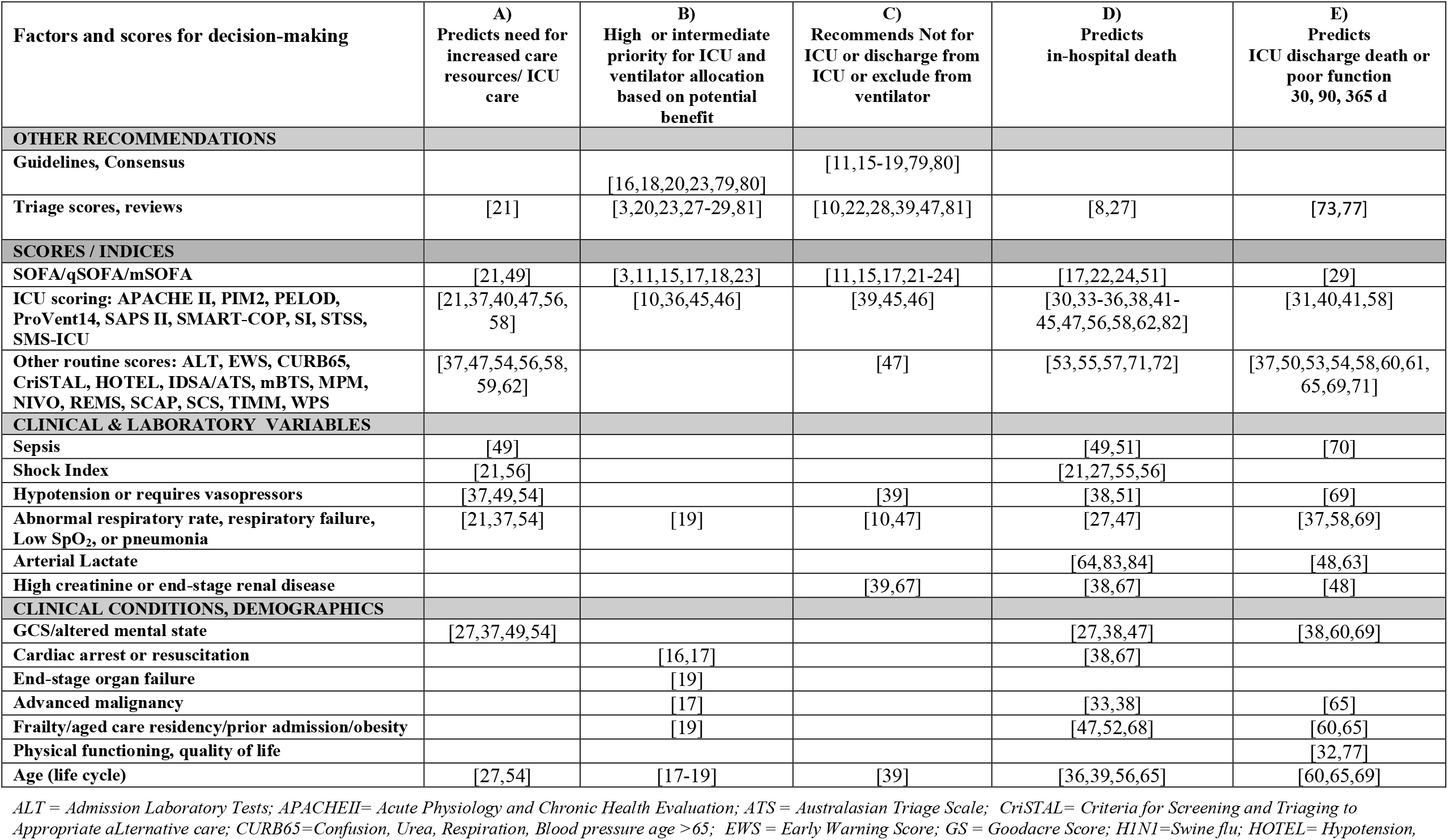
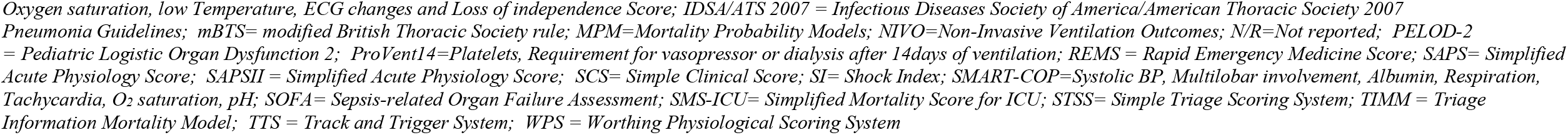
Evidence-based variables and scores for decision-making about allocation of ICU care and ventilators based on predicted outcomes (during pandemics and routine care)

| Factors and scores for decision-making | A)<br>Predicts need for<br>increased care<br>resources/ ICU<br>care | B)<br>High or intermediate<br>priority for ICU and<br>ventilator allocation<br>based on potential<br>benefit | C)<br>Recommends Not for<br>ICU or discharge from<br>ICU or exclude from<br>ventilator | D)<br>Predicts<br>in-hospital death | E)<br>Predicts<br>ICU discharge death or<br>poor function<br>30, 90, 365 d |
| --- | --- | --- | --- | --- | --- |
| **OTHER RECOMMENDATIONS** |  |  |  |  |  |
| Guidelines, Consensus |  | [16,18,20,23,79,80] | [11,15-19,79,80] |  |  |
| Triage scores, reviews | [21] | [3,20,23,27-29,81] | [10,22,28,39,47,81] | [8,27] | [73,77] |
| **SCORES / INDICES** |  |  |  |  |  |
| SOFA/qSOFA/mSOFA | [21,49] | [3,11,15,17,18,23] | [11,15,17,21-24] | [17,22,24,51] | [29] |
| ICU scoring: APACHE II, PIM2, PELOD, ProVent14, SAPS II, SMART-COP, SI, STSS, SMS-ICU | [21,37,40,47,56,58] | [10,36,45,46] | [39,45,46] | [30,33-36,38,41-45,47,56,58,62,82] | [31,40,41,58] |
| Other routine scores: ALT, EWS, CURB65, CriSTAL, HOTEL, IDSA/ATS, mBTS, MPM, NIVO, REMS, SCAP, SCS, TIMM, WPS | [37,47,54,56,58,59,62] |  | [47] | [53,55,57,71,72] | [37,50,53,54,58,60,61,65,69,71] |
| **CLINICAL & LABORATORY VARIABLES** |  |  |  |  |  |
| Sepsis | [49] |  |  | [49,51] | [70] |
| Shock Index | [21,56] |  |  | [21,27,55,56] |  |
| Hypotension or requires vasopressors | [37,49,54] |  | [39] | [38,51] | [69] |
| Abnormal respiratory rate, respiratory failure, Low SpO<sub>2</sub>, or pneumonia | [21,37,54] | [19] | [10,47] | [27,47] | [37,58,69] |
| Arterial Lactate |  |  |  | [64,83,84] | [48,63] |
| High creatinine or end-stage renal disease |  |  | [39,67] | [38,67] | [48] |
| **CLINICAL CONDITIONS, DEMOGRAPHICS** |  |  |  |  |  |
| GCS/altered mental state | [27,37,49,54] |  |  | [27,38,47] | [38,60,69] |
| Cardiac arrest or resuscitation |  | [16,17] |  | [38,67] |  |
| End-stage organ failure |  | [19] |  |  |  |
| Advanced malignancy |  | [17] |  | [33,38] | [65] |
| Frailty/aged care residency/prior admission/obesity |  | [19] |  | [47,52,68] | [60,65] |
| Physical functioning, quality of life |  |  |  |  | [32,77] |
| Age (life cycle) | [27,54] | [17-19] | [39] | [36,39,56,65] | [60,65,69] |

ALT = Admission Laboratory Tests; APACHEII= Acute Physiology and Chronic Health Evaluation; ATS = Australasian Triage Scale; CriSTAL= Criteria for Screening and Triaging to Appropriate aLternative care; CURB65=Confusion, Urea, Respiration, Blood pressure age >65; EWS = Early Warning Score; GS = Goodacre Score; H1N1=Swine flu; HOTEL= Hypotension,
*Oxygen saturation, low Temperature, ECG changes and Loss of independence Score; IDSA/ATS 2007 = Infectious Diseases Society of America/American Thoracic Society 2007 Pneumonia Guidelines; mBTS= modified British Thoracic Society rule; MPM=Mortality Probability Models; NIVO=Non-Invasive Ventilation Outcomes; N/R=Not reported; PELOD-2 = Pediatric Logistic Organ Dysfunction 2; ProVent14=Platelets, Requirement for vasopressor or dialysis after 14days of ventilation; REMS = Rapid Emergency Medicine Score; SAPS= Simplified Acute Physiology Score; SAPSII = Simplified Acute Physiology Score; SCS= Simple Clinical Score; SI= Shock Index; SMART-COP=Systolic BP, Multilobar involvement, Albumin, Respiration, Tachycardia, O<sub>2</sub> saturation, pH; SOFA= Sepsis-related Organ Failure Assessment; SMS-ICU= Simplified Mortality Score for ICU; STSS= Simple Triage Scoring System; TIMM = Triage Information Mortality Model; TTS = Track and Trigger System; WPS = Worthing Physiological Scoring System*

The Sequential Organ Failure Assessment (SOFA) score and its variants was the most widely used (or reported) for both ICU admission and discharge criteria, as well as to recommend ventilator allocation or removal.

Patients who stand to benefit the most from ICU admission typically suffer from a critically severe, treatable and potentially reversible deterioration of health. ICU treatment should also be consistent with the values and preferences of the patient [13,74]. When patients’ are deteriorating despite ongoing ICU care, withdrawal of life-sustaining therapies, and transfer to ward and palliative care has to be considered. This process is known as ‘reverse triage’ [75] and variables to facilitate these decisions are listed in Table 1, column C. Withholding or withdrawing treatments must include discussions with the patient (if possible) and their family. Ideally during pandemic triage the possibility of future deterioration and need to discharge from ICU later should already be discussed on admission to ICU.

Multiple studies investigated predictors for mortality in the ensuing weeks and months after ED or ICU admission to inform decision-making, mostly in non-pandemic situations (Table 1, columns D and E). These prediction models can assist clinicians and patients in the decision-making about the appropriateness of ICU care by providing information about the expected recovery (or likely downward trajectory) following ICU admission and/or ventilator treatment [39,76,77].

### Variables and prognostic factors used for decision-making in pandemic situations

Amongst the sixteen pandemic-related publications, several expert consensus documents outlined factors that inform the allocation of ICU care and ventilator treatment (Table 2). The usual criteria for requiring critical care interventions applied such as refractory hypoxaemia, respiratory acidosis, evidence of impending respiratory failure, shock, requiring, vasopressor or inotropic support, decreased urinary output, evidence of organ failure, and altered level of consciousness.

**Table 2.**
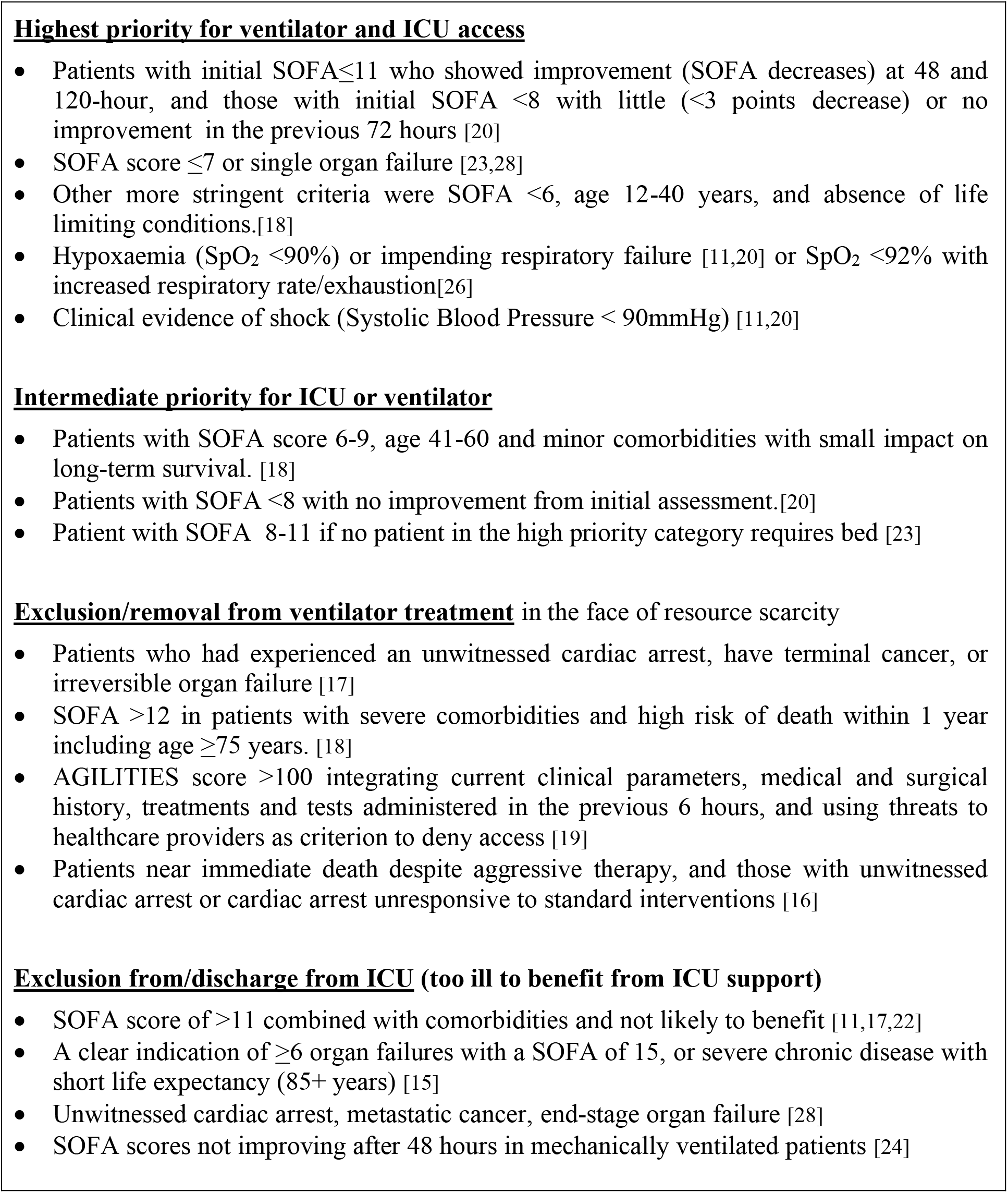
Summary of consensus/taskforce/algorithm recommendations for rationing ICU care and ventilators during pandemics according to patient status and response to treatment.

The SOFA, qSOFA, and mSOFA score cut-offs were used to either determine priority for ICU or ventilator access or removal from a ventilator and/or discharge from ICU in futile situations. Likewise, the AGILITIES score and Simple Triage Scoring (STSS) for adults, the Pediatric Logistic Organ Dysfunction 2 (PELOD) triage score, the COVID-19 clinical risk score, and the Community Assessment Tools (CATs) criteria for adults and children were used to prioritise ICU admission or recommend ICU discharge due to a lack of benefit.

Age was a variable in six of the 15 predictive approaches for pandemic situations [8,18,19,25,27,78]. Two studies outlined criteria for patients who do not require organ support or ICU because they are below the critical illness threshold [11,23].

Table S2.1 in Supplement 2 lists specific details of some tools used for decision-making about ICU admission and ventilator allocation during pandemics along with their reported accuracy (AUROC, OR or sensitivity/specificity). Recommendations in patients who warrant high priority access to ICU care but have a poor prognosis were ambiguous. The COVID-19 clinical risk score predicted either need for critical care, need for ventilator or the risk of in-hospital death for patients with COVID-19, based on age, abnormal chest radiography, haemoptysis, dyspnoea, unconsciousness and history of cancer [25]. Another triage tool also had composite outcomes for patients with a score of ≥3 (including age, respiratory rate, oxygen saturation, shock index and altered mental status) [27]. The CATs tool was based on 5 respiratory criteria and evidence of shock and altered level of consciousness to predict the need for mechanical ventilation or ICU admission or the risk of death but did not indicate when to set a threshold for choosing a care pathway [26].

### Prognostic tools used in non-crisis situations

Our study also explored whether triage algorithms/factors used for ICU, emergency departments or in hospital wards in non-crisis situations (Supplement 2, Table S2.2), could add value to the above pandemic recommendations. The majority of these prognostic indicators were derived from large patient population studies and predicted in-hospital or post-discharge mortality. Only four indicators were based on expert consensus, of which three [29,49,51] used SOFA or qSOFA for predicting outcomes, while a ward-based rule to predict mortality used the CURB65 score [69]. The tools applied predominantly to adult patients in routine ICU care (18 studies) or patients being assessed in emergency departments (21 studies), with only a few (5 studies) used in routine ward care.

Unlike pandemic tools, which focus on acute organ failure, many routine care decision-making algorithms rely more on patient history of chronic illness [34,38,41,46,52,60,61,65,66,68], admission type (emergency, medical, elective surgery, non-trauma) [30,31,35,38,39,41,65], and age [38,39,46,54,57,59,60,62,65,68,70,71].

### Triage algorithms, scores and tools used in non-crisis ICU care

Instruments for decision-making about ICU triage this category also included SOFA, and qSOFA, but a diverse collection was a: Simplified Mortality Score for the ICU (SMS-ICU), ProVent 14 score, Simplified Acute Physiology Score (SAPSS II, SAPS III), SMART –COP score, Mortality Probability Models (MPM), and Acute Physiology and Chronic Evaluation (APACHE II, III) and for children the Revised Paediatric Index of Mortality and PELOD-2.

ICU-based algorithms relied predominantly on laboratory variables or acute treatments such as those for sepsis or respiratory failure [35,37,39,41-43,46] and two relied solely on a single biomarker cut-off: Secretoneurin [44] and Procalcitonin respectively [45]. Five of the 18 articles on routine care included algorithms to predict clinical deterioration with a need for ICU admission [36,37,40,46,49]. Only one article included a tool to predict the need for vasopressor treatment and respiratory support [37]. Two articles provided information on patients who are unlikely to benefit from (ongoing) ICU treatment [39,46].

### Emergency Department algorithms used in non-crisis situations

Emergency department decision-making algorithms combined laboratory tests and clinical history or examination: Severe community acquired pneumonia score (SCAP), Infectious Diseases Society of America/American Thoracic Society IDSA/ATS), the Shock Index (SI), Mortality in Emergency Department Sepsis (MEDS) score, Simple Clinical Score (SCS), Emergency Severity Index (ESI), Triage Information Mortality model (TIMM), and Criteria for screening and triaging to appropriate alternative care (CriSTAL). Two studies predicted in-patient mortality based only on frailty syndrome [52,68], and six studies based mortality predictions purely on laboratory test results [48,50,63,64,66,67].

Six of the 21 studies among patients in the emergency department predicted the need for potential transfer to ICU based on clinical and laboratory variables [54,56,58,61,62,66]. The other studies predicted mortality at different time points. One study focused on decision-making about ICU admission in patients on chronic dialysis [67]. Two studies provided recommended a score cut-off for referral to palliative care [61,65].

### Ward-based triage used in non-crisis situations

Five scoring systems predicted in-hospital and 30-day mortality among ward-based patients (Simple Clinical Score, HOTEL, CURB65, NIVO, and Mortality Predictive Model for Children (MPMC) based on a combination of clinical criteria and laboratory test results [47,69-72]. One scoring system predicted the need for non-invasive ventilation in ward-based patients with COPD and was recommended for setting a ceiling of treatment [47].

### Summary of variables and prognostic factors used for decision-making in non-pandemic situations

Table S2.2 in Supplement 2 gives an overview of variables predicting poor patient outcome for decision-making about ICU admission (IDSA/ATS, ESI) or discharge in non-pandemic situations presented for ICU, ED and ward care. Several tools including SAPS II, APACHE II/III, SOFA) predicted in-hospital mortality risk. Results from the validation of Simple Clinical Score, CriSTAL tool, and Shock index, indicated a good predictive value to identify people who will require ICU admission, palliative care or will die in the short term post-discharge.

### Using combined variables and prognostic factors for decision-making in pandemic situations

A simplified example of triage recommendations for pandemic times based on predicted prognosis is illustrated in Figure 2. The parameters and accompanying cut-off points are extracted from the comprehensive factors used in both pandemic and routine care shown in Supplement 2. Elements from this catalogue of triage criteria could be used to design locally relevant triage tools.

In this priority setting scenario for pandemics, two types of patients are unambiguously outside eligibility thresholds and therefore excluded from access to critical care resources: patients in group 1 who are ‘too healthy’ and patients in group 5 who are ‘too sick’ (Figure 2). For other severity profiles (patients in groups 2 to 5 with increasingly poorer prognosis) some parameters may be unknown at triage; a suite of alternatives are listed to assist with decision-making. Periodic reassessment at 48, 96 and 120 hours is recommended [11,20,24,28,38] to determine the need to discharge to ward due to improvement, escalate treatment to ICU, or withdraw ICU treatment and refer for palliative care as indicated by the arrows between all groups.

**<Figure 2. Sample prioritisation criteria to determine access or exclusion from critical care services during pandemics using combinations of crisis and non-crisis algorithms> about here**

The concept of a waiting list may not apply under normal circumstances, but in overwhelmed health systems, people with characteristics in group 4 may have to be managed on the ward until an ICU bed becomes available and no patient with a higher priority profile is competing for an ICU bed.

Patients who are deemed to be beyond salvageable –group 5 with the poorest prognosis– usually have already experienced a catastrophic event like a cardiac arrest and/or are unconscious and/or are refractory to vasopressors and/or need or have been on mechanical ventilation for 14+ days and/or have documented advanced chronic illness/frailty/age. The general recommendation is not to use scarce resources on these patients during a public health crisis. Importantly, no decision should be based on single parameters or undesirable individual characteristics.

### Ethical and resource considerations in crisis decision-making

A number of publications identified in this scoping review explored social, ethical, and political considerations when making decisions about patients’ access to ICU and ventilator treatment (Table 3). Key principles included institutional and public transparency about the decision-making process, consultation with interdisciplinary groups, and establishing local partnerships for implementation of decision-making processes in a local context. Studies varied in their approach to including patients’ illness severity scores in the decision-making process, with some supporting their inclusion due to objectivity and validity for patient groups [11,18], and others recommending against their use in isolation to predict individual patient outcomes [80].

**Table 3.** Ethical, legal, practical and clinical considerations when allocating ICU care ventilator treatment.

| What to consider in ensuring fair rationing of resources |
| --- |
| ***Ethical, legal and practical considerations***<br>• Make triage policy and rationing criteria transparent to staff and the public to ensure understanding of the reasons for access restrictions [2,15,79,80]<br>• Establish local/regional partnerships to effectively manage resource shortages and triage pathways [2,15,75,80]<br>• When resources cannot be maintained, unequal treatment may be justified and choice determined on the basis of medical need and likely benefit[12] by using a proportionately lower amount of resources [81]<br>• People making decisions about resource allocation should not be the treating clinicians but preferably a central triage committee of senior clinicians and legal officers [15,79,86]<br>• First come first served or lottery/ random assignment if two patients with same level of risk present at the same time [81]<br>• Decision-makers should not be aware of patients' identity [19]<br>• Revise existing laws and develop liability protection for clinicians using scoring systems for decision-making about allocation of scarce resources [15,29]<br>• Priority to be given to those who have had less opportunity to live their lives [2,29]<br>• Align patients values and preferences with decisions to admit or remain in ICU [87]<br>• Build capacity for alternatives to ICU care before life-sustaining resources become limited and provide alternative options such as high quality palliative care [15]<br>• Ongoing update of the triage process as new evidence of disaster experiences, research, planning, and modelling becomes available [15]<br>***Clinical decisions***<br>• Educate lay people to better understand goals of care so they can be partners in a shared decision-making process [74,88]<br>• Use best available objective parameters to determine patients' risk profile [15], and allocate patients to waiting lists or immediate resource access [15]<br>• Triage patients to higher level of care if they have the greatest medical need and higher chances of survival [20,81]<br>• Re-assess response to ventilator treatment and other therapies periodically, to see if patients are recovering or if they are deteriorating to confirm that the eligibility criteria are still met (i.e. risk of death, lack of improvement of SOFA after 48 hours) [19,24,81]<br>• If the patient does not deteriorate, commit the ventilator for 10-12 days[3]. For patients on mechanical ventilation for 14+ days, clinicians should have a discussion with the family about the poor prognosis and goals of care [39]<br>• Refer to palliative care from ED if risk of death is very high (i.e. prior CPR or cardiac arrest)[16,17,29]<br>• Discharge patients from ICU who have an increased mortality risk or are unlikely to benefit from ICU treatment [15,17,22,24]<br>• Apply objective criteria, set ceilings of treatment, discuss with families a referral to palliative care, and remove those from ventilator treatment who no longer meet objective criteria for benefit [10,18,19] |
| What to avoid when deciding on ICU/ventilator allocation |
| • Considering older age as single criterion for exclusion from higher level of care [17,29,36,65]<br>• Basing decisions on gender, race, gender, sexual orientation, religion, disability, social status, personal connections, wealth, citizenship, insurance status [19,80]<br>• Using obesity to inform prognosis in isolation from other risk factors [18]<br>• Prioritising healthcare workers [17],people with dependent parents or children [81]<br>• Allocating a ventilator to a pregnant woman to save a non-viable foetus (<24 weeks gestation) although appropriate to save the woman [19]<br>• Triaging people with a single condition to ICU ahead of those with comorbidities [17]<br>• Using the number of medical conditions rather than their severity to make decisions [38]<br>• Using Shock Index in isolation to rule out triage to ICU [56]<br>• Offering short trials of ICU to patients who are not eligible (low illness severity, minor comorbidities, young age) [18,80]<br>• Offering ICU beds to patients whose authentic values or informed treatment preferences are not consistent with ICU treatment/admission [13,74] |

Guidelines warned against making judgments about the worth of individuals, and unintentionally discriminating population subgroups such as the elderly or obese, or people with certain conditions as well as specific ethnic groups or people in non-health occupations. However, some publications outlined that older age is associated with a higher risk of death [19,21,25] or flagged a natural shorter life expectancy as an additional reason for excluding patients from critical care [15], and indicated that among young people most years of life could potentially be saved [18]. Giving preferential treatment to population subgroups such as those with dependent children [81], caregivers of elderly [81] or frontline pandemic health workers [17] were discouraged by some and promoted by others [80,85]. Likewise, discrepancies were found about the recommendation to (not) involve treating clinicians in the decision-making process, with some studies supporting the exclusion of the treating clinicians [15,86] while others recommended that the senior treating clinician should lead the decision-making process [80].

## Discussion

This scoping review identified key principles of decision-making about allocation of intensive care beds and ventilator treatment during pandemics including the need for a local expert committee of decision-makers, use of best available objective clinical criteria, careful steering from unintentional discrimination of vulnerable groups, consistent application of rules, transparency, and ethical justification of service limitations. Our review identified a range of tools to support the decision-making process and outlined their characteristics (62 publications on guidelines, frameworks, algorithms, laboratory parameters and predictive tools).

In addition, we systematically collated scores and algorithms that can be used for triage decision-making in routine care (in non-pandemic situations) in the emergency department, on the wards, or in the ICU. Some of the identified tools were derived from influenza pandemics and non-respiratory disease public health emergencies, yet they can be extrapolated to other public health emergencies including COVID-19.

### How do the results fit in with previous research and policies?

It is generally accepted that triage protocols should only be activated when resource scarcity is imminent [21,46]; be locally relevant through a committee of expert decision-makers; make best use of relevant objective criteria; be carefully steered from unintentionally discriminating vulnerable groups; consistently apply agreed rules; and be publicly transparent and ethically justifiable. ICU admission criteria vary from country to country [89] and can change overtime as technology advances, but generally rely on expert assessment of the patient’s illness severity, the health system culture, resources, patient preference, and ethical considerations. Admission is considered appropriate from the medical perspective, for those who are likely to require mechanical ventilation, or support for single or multiple organ failure [89]. However, when resources are overwhelmed by a surge in number of cases requiring escalation of care, these criteria need to vary towards “crisis standards of care”[90].

It has been suggested that vital signs in critically ill patients can inform triage decisions during pandemics [27]. Quick and simplified scores e.g. qSOFA vs. SOFA) have been developed to predict sepsis mortality. However, there might be trade-offs between the simplicity of a score and its sensitivity and specificity. Using such scoring systems to deny access to care is controversial. [91]Complex algorithms with multiple variables increase the burden of data collection without necessarily increasing the predictive ability (e.g. APACHE), but some scores with multiple variables have been shown to increase predictive ability (SIRS and NEWS). The appeal of some relatively simple predictive tools (mSOFA, qSOFA, SCS, CriSTAL, AGILITIES) is that they do not require additional testing, although some clinicians warn against the use of population-based algorithms in isolation to guide decisions for individuals [80].

### No consensus on ethical and social considerations

We found mixed support for some of the subjective criteria in the expert consensus and triage publications. Fears of discrimination of elderly, functionally impaired, cognitively impaired, obese or immunosuppressed patients when allocating resources have been publicly expressed [18]. Under the “life cycle principle” younger patients receive priority because they have had the least opportunity to live through stages of life [18]. A different ethical principle to make allocation decisions is the “maximizing life-years” which takes into account a patient’s life expectancy based on age, co-morbidities and other factors (hence prioritising the young) [29]. The principle behind giving healthcare workers priority [85] is a ‘multiplier effect’. When healthcare workers recover they can contribute to saving the lives of many others. Although this principle may have some ethical validity, it did not receive much support in the consensus statements.

### Implementation strategies to manage clinician burden and public dissatisfaction

In overwhelmed health systems, many people with poorer prognosis may be diverted to palliative care, thus increasing demand for accelerated staff training in the communication with patients and families about end-of-life care [92]. Some publications suggested involvement of an external expert committee of senior clinicians to make decisions about ceilings of care in order to reduce the burden on the treating clinicians [15,19,86]. A legal framework needs to be in place to protect healthcare workers from litigation if they allocate limited resources in accordance with ethical guidelines [15,93,94].

Triage aims to maximise positive health outcomes for the largest possible number of patients. Triage protocols can have negative consequences for patients who are already hospitalised or treated in ICU for conditions not related to a pandemic (e.g. stroke or myocardial infarction) who would not have been denied access under normal conditions [23]. Those admitted to ICU, should be reassessed at 48 and up to 120 hours to determine ongoing eligibility for ICU resources [21,24] or to discharge them to palliative care. While an early, gradual and personalised approach to prognostic disclosure in routine practice is recommended in terminal illness [95], this may not be possible in mass emergencies. Early prognostic disclosure may be necessary during pandemics and other public health emergencies, and “advance care planning” will have to be expedited at the time of admission. Patient and family involvement in treatment decisions may be limited by hospital policies concerned with service capacity and healthcare worker safety. However, when possible, recognising triggers for early palliative care referral and/or treatment withdrawal [96] and adhering to patient preferences [97] should be integral to management policies.

It has been recommended to invite input from members of at-risk groups or their caregivers into algorithms to determine access to ICU and ventilators during pandemics [18,93,98]. However, this may not work in all cultures, and time pressure will likely be prohibitive of consultation with all stake holders. Unfortunately, efficient allocation of ventilators may unintentionally further increase social inequalities [99]. Supplementary strategies to build trust during a pandemic include public transparency on the objective decision-making framework [18,98] and disseminating information about decision-making frameworks to the public.

### Strengths and limitations of this review

We believe that this scoping review provides a useful resource for decision-making about ICU and ventilator allocation during pandemics. This is a discussion starter and can inform objective guidelines beyond the guiding principles of preparedness, organisational management for resource allocation, expanded scope of practice, equity and social justice currently published [3,79,100-103]. Prognostic indicators and other decision tools presented have been based on a multitude on criteria, which can be combined to create locally-relevant triage algorithms. This is a discussion starter and can inform objective guidelines beyond the guiding principles of equity and social justice currently published.

We did not conduct risk of bias assessment of included studies as the purpose of this scoping review was to collate a wide range of risk prediction and decision tools, which will have to be adapted to local settings. We excluded some validation studies from low-income countries [104] which showed good predictive ability of the combined variables as there was the chance of lesser generalisability of their patient population to health systems in industrialised nations –the focus of our study.

## Conclusions

The catalogue of resources we assembled provides guidance on variables used to prioritise patients for critical care in the face of scarce life-sustaining resources. Patients’ clinical or demographic characteristics alone and rigid triage systems are not the preferred way of allocating resources in a constrained healthcare system. The patient perspective needs to be taken into account. Discrimination against certain population groups must be avoided at every level of disease severity. A combination of variables used in prognostic scores (based on chronic and acute risk factors) and other decision tools presented here can be combined to create locally-relevant triage algorithms to assist decisions about ICU admission and discharge and/or access to ventilator treatments during a pandemic. This unique resource will help service managers and clinicians with the emotional and ethical burden of having to select some patients over others for life-sustaining treatments. More importantly objective guidelines will provide transparency about rationing resources to the patients and communities they serve.

## Data Availability

All data available is reported in tables or text. Any requests can be addressed to the corresponding author

## ONLINE SUPPLEMENTS 1 and 2

**Supplement 1. Table S1.1.**
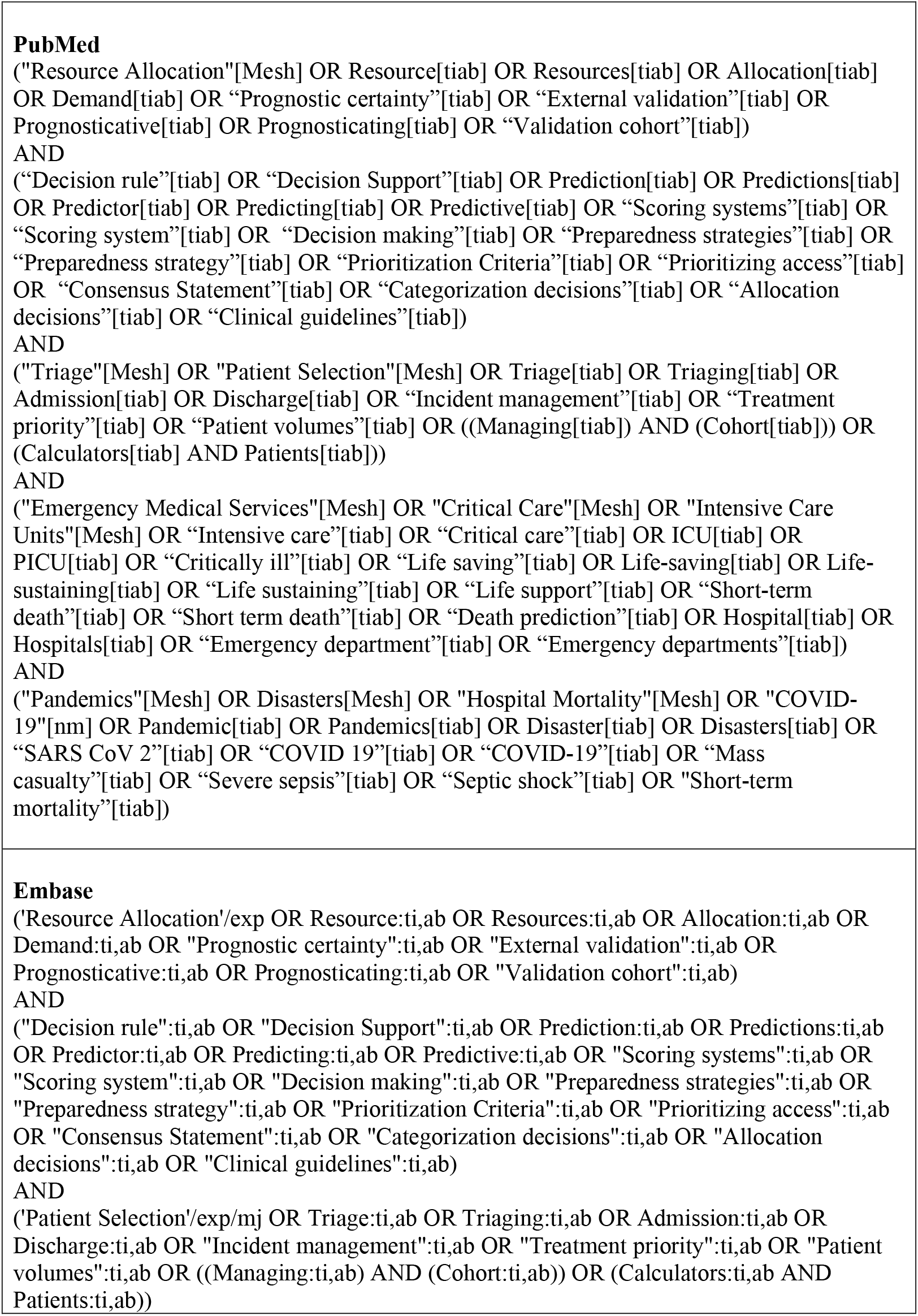
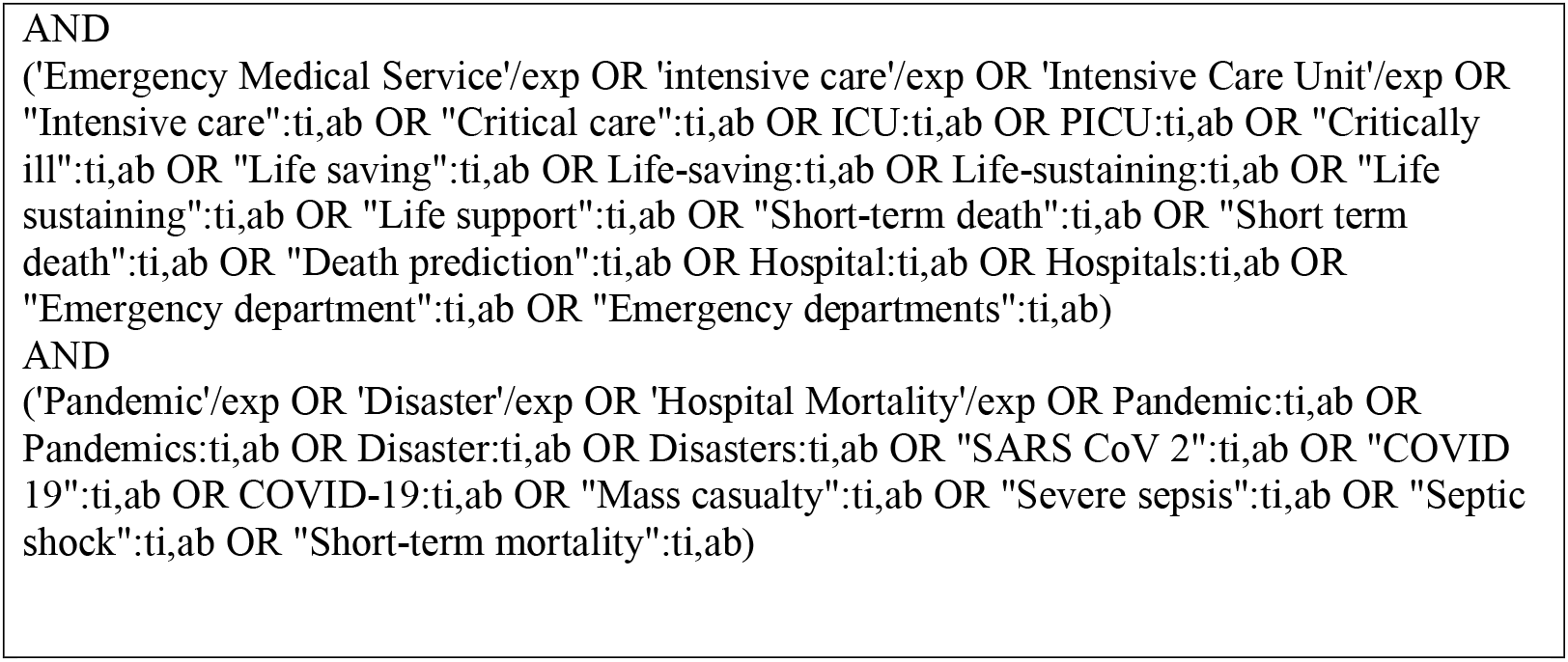
Detailed Search strategy Searches run 14/05/2020:

**Supplement 1, Table S1.2.** Reasons for exclusion after full text screening N= 70.

| **No objective parameters** | **N=9** |
| --- | --- |
| Barrera, R., et al. (2001). "Accuracy of predictions of survival at admission to the intensive care unit." *J Crit Care* 16(1): 32-35. |  |
| Burkle, F. M., Jr. (2006). "Population-based triage management in response to surge-capacity requirements during a large-scale bioevent disaster." *Acad Emerg Med* 13(11): 1118-1129. |  |
| Christian, M. D., et al. (2014). "Triage: care of the critically ill and injured during pandemics and disasters: CHEST consensus statement." *Chest* 146(4 Suppl): e61S-74S. |  |
| Dadoun, A., et al. (2017). "Invoking the "expectant" triage category: Can we make the paradigm shift?" *Am J Disaster Med* 12(3): 167-172. |  |
| Einav, S., et al. (2014). "Surge capacity logistics: care of the critically ill and injured during pandemics and disasters: CHEST consensus statement." *Chest* 146(4 Suppl): e17S-43S. |  |
| Fink, S. L. (2010). "Worst case: rethinking tertiary triage protocols in pandemics and other health emergencies." *Crit Care* 14(1): 103. |  |
| Godfrey, G., et al. (2012). "Treatment limitations at admission to intensive care units in Australia and New Zealand: prevalence, outcomes, and resource use*." *Crit Care Med* 40(7): 2082-2089. |  |
| Prekker ME, et al., (2020) Regional planning for extracorporeal membrane oxygenation allocation during Coronavirus disease 2019. *Chest* 158(2):603-607. |  |
| Ravikumar N, et al (2020) Novel Coronavirus (2019-nCoV) Infection: Part I – Preparedness and Management in the pediatric intensive care unit in resource-limited settings. *Indian Pediatrics* April 15, p324-334 |  |
| **No validation or regression** | **N=8** |
| Antommaria, A. H., et al. (2020). "Ventilator Triage Policies During the COVID-19 Pandemic at U.S. Hospitals Associated With Members of the Association of Bioethics Program Directors." *Ann Intern Med*. |  |
| Baysan, M., et al. (2020). "The Added Value of Lactate and Lactate Clearance in Prediction of In-Hospital Mortality in Critically Ill Patients With Sepsis." *Crit Care Explor* 2(3): e0087. |  |
| Cowen, M. E., et al. (2014). "Implementation of a mortality prediction rule for real-time decision making: feasibility and validity." *J Hosp Med* 9(11): 720-726 |  |
| Dziadzko, M. A., et al. (2018). "Multicenter derivation and validation of an early warning score for acute respiratory failure or death in the hospital." *Crit Care* 22(1): 286. |  |
| Hahn, R., et al. (2009). "Use of pediatric risk of mortality (PRISM) II score as a tool for critical care triage during a pandemic." *Critical Care Medicine* 37(12): A327. |  |
| Hick J. L., et al. (2007) Clinical review: Allocating ventilators during large-scale disasters - problems, planning, and process |  |
| Thomas, D. R., et al. (2005). "The relationship of functional status, nutritional assessment, and severity of illness to in-hospital mortality." *J Nutr Health Aging* 9(3): 169-175 |  |
| Thakur, M., et al. (2016). "Intensive Care Admissions in Pregnancy: Analysis of a Level of Support Scoring System." *Matern Child Health J* 20(1): 106-113. |  |
| **Abstract/Poster Presentation-insufficient information** | **N=9** |
| Acton, C., et al. (2016). "Ten patients, one ventilator: How to best allocate critical care resources during mass disaster." *Canadian Journal of Emergency Medicine* 18: S79. |  |
| Adeniji, K., et al. (2010). "The simple triage scoring system (STSS) successfully predicts mortality and critical care resource utilisation in H1N1 pandemic flu." *Intensive Care Medicine* 36: S286. |  |
| Allareddy, V., et al. (2018). "Use of ecmo and associated outcomes in children hospitalized for sepsis in the United States." *Critical Care Medicine* 46: 744. |  |
| Aluvaala, M. J., et al. (2018). "Predicting in-hospital mortality using routine data to guide decision making for essential neonatal care." *American Journal of Tropical Medicine and Hygiene* 99(4): 450. |  |
| Flaatten, H. (2009). "ICU triage: An overview." *Acta Anaesthesiologica Scandinavica* 53: 17-18. |  |
| Marriott, D., et al. (2012) "To admit or not to admit? The suitability of critical care admission criteria." *Crit Care* 16, P511 |  |
| Nobre de Jesus et al (2019) "Clinical outcome prediction after admission in intermediate level critical care unit:role of severity scores." *Critical Care* 23(Suppl 2):72, poster 466 |  |
| Soliman, I. W., et al. (2017). "Long-term prognostication in the ICU: A systematic review into available prediction models." *Intensive Care Medicine Experimental* 5(2). |  |
| Soong, J. T. Y., et al. (2017). "Frailty syndromes coded within secondary user service(SUS) data predict inpatient mortality and long length of stay." *Acute Medicine* 16(3): 131-132. |  |
| **Ethical/resource/preparedness focus** | **N=11** |
| Australasian College of Emergency Medicine. "Clinical Guidelines for the management of COVID-19 in Australasian Emergency Departments". Melbourne VIC: ACEM, 2020, p. 112. |  |
| Antommaria, A. H., et al. (2010). "Critical appraisal of: triaging pediatric critical care resources during a pandemic: ethical and medical considerations." *Pediatr Crit Care Med* 11(3): 396-400. |  |
| Antommara, A. H., et al. (2011). "Ethical issues in pediatric emergency mass critical care." *Pediatr Crit Care Med* 12(6 Suppl): S163-168. |  |
| Barnett, D. J., et al. (2009). Resource allocation on the frontlines of public health preparedness and response: report of a summit on legal and ethical issues. *Public Health Rep* 124(2): 295-303. |  |
| Biddison, L. D., et al. (2014). "Ethical considerations: care of the critically ill and injured during pandemics and disasters: CHEST consensus statement." *Chest* 146(4 Suppl): e145S-155S. |  |
| Challen K, et al., (2007) "Clinical review: mass casualty triage – pandemic influenza and critical care". *Critical Care* 11:212, 6pp |  |
| Farrell, T. W., et al. (2020). "AGS Position Statement: Resource Allocation Strategies and Age-Related Considerations in the COVID-19 Era and Beyond." *J Am Geriatr Soc*. |  |
| Flaatten, H., et al. (2017). "The status of intensive care medicine research and a future agenda for very old patients in the ICU." *Intensive Care Med* 43(9): 1319-1328. |  |
| Ghanbari, V., et al. (2019). "Ethical prioritization of patients during disaster triage: A systematic review of current evidence." *Int Emerg Nurs* 43: 126-132. |  |
| Kanter, R. K., & Cooper, A. (2009). Mass critical care: pediatric considerations in extending and rationing care in public health emergencies. *Disaster medicine and public health preparedness*, 3(S2), S166-S171. |  |
| Maves RC, et al., (2020) "Triage of scarce critical resources in COVID-19. An implementation guide for regional allocation". *Chest* 158(1):212-225 |  |
| **Modelling study/no primary data** | **N=11** |
| Abir, M., et al. (2013). "Design of a model to predict surge capacity bottlenecks for burn mass casualties at a large academic medical center." *Prehosp Disaster Med* 28(1): 23-32. |  |
| Amram, O., et al. (2012). "A web-based model to support patient-to-hospital allocation in mass casualty incidents." *J Trauma Acute Care Surg* 72(5): 1323-1328. |  |
| Bolt, S. and R. Sparks (2013). "Detecting and diagnosing hotspots for the enhanced management of hospital Emergency Departments in Queensland, Australia." *BMC Med Inform Decis Mak* 13: 132. |  |
| Enfield, K., et al. (2011). "Limited utility of SOFA and APACHE II prediction models for ICU triage in pandemic influenza." *Chest* 140(4). |  |
| Ercole, A., et al. (2009). "Modelling the impact of an influenza A/H1N1 pandemic on critical care demand from early pathogenicity data: the case for sentinel reporting." *Anaesthesia* 64(9): 937-941. |  |
| Gall, C., et al. (2016). "Pediatric Triage in a Severe Pandemic: Maximizing Survival by Establishing Triage Thresholds." *Crit Care Med* 44(9): 1762-1768. |  |
| Kanter, R. K. (2015). "Would triage predictors perform better than first-come, first-served in pandemic ventilator allocation?" *Chest* 147(1): 102-108. |  |
| Nap, R. E., et al. (2007). "Pandemic influenza and hospital resources." *Emerg Infect Dis* 13(11): 1714-1719. |  |
| Toltzis P, Soto-Campos G, Kuhn E, Wetzel R. A pediatric triage scheme to guide resource allocation in a mass casualty. *Critical Care Medicine*. 2013;41(12):A148. |  |
| Steinberg DO, et al. (2020) Critical review: COVID-19 calculators during extreme resource –limited situations. *Emerg Medicine Pract* 22(5):5pp |  |
| Toltzis P, Soto-Campos G, Shelton C, Kuhn EM, Hahn R, Kanter RK, et al. Evidence-Based Pediatric Outcome Predictors to Guide the Allocation of Critical Care Resources in a Mass Casualty Event. *Pediatr Crit Care Med*. 2015 Sep;16(7):e207-16. |  |
| **Non-Western health system** | **N=6** |
| Alam A, Gupta S. Lactate Measurements and Their Association With Mortality in Pediatric Severe Sepsis in India: Evidence That 6-Hour Level Performs Best. *J Intensive Care Med*. 2020 |  |
| Arshad A, Ayaz A, Haroon MA, Jamil B, Hussain E. Frequency and Cause of Readmissions in Sepsis Patients Presenting to a Tertiary Care Hospital in a Low Middle Income Country. *Crit Care Explor*. 2020 Feb;2(2):e0080. |  |
| Santana AR, Amorim FF, Menezes BM, Soares FB, Araújo FVB, De Souza JL, et al. Serum arterial lactate at the time of admission as a predictor of mortality in patients admitted with severe sepsis and septic shock to an ICU. *Critical Care*. 2013;17:P47. |  |
| Ytzhak A, Sagi R, Bader T, Assa A, Farfel A, Merin O, et al. Pediatric ventilation in a disaster: clinical and ethical decision making. *Crit Care Med*. 2012 Feb;40(2):603-7. |  |
| Dumas, G., et al. (2019). "Mottling score is a strong predictor of 14-day mortality in septic patients whatever vasopressor doses and other tissue perfusion parameters." *Crit Care* 23(1): 211. |  |
| Yusoff SJ, Zakaria MI, Mohamed FL, Bujang MA, Lakshmanan S, Asaari AH. Value of Shock Index in prognosticating the short-term outcome of death for patients presenting with severe sepsis and septic shock in the emergency department. *Med J Malaysia*. 2012 Aug;67(4):406-11. |  |
| **Other disease/treatment specific** | **N=4** |
| Barbaro, R. P., et al. (2016). "Development and validation of the pediatric risk estimate score for children using extracorporeal respiratory support (Ped-RESCUERS)." *Intensive Care Med* 42(5): 879-888 |  |
| Bozcuk H, Koyuncu E, Yildiz M, Samur M, Ozdogan M, Artaç M, et al. A simple and accurate prediction model to estimate the intrahospital mortality risk of hospitalised cancer patients. *Int J Clin Pract*. 2004 Nov;58(11):1014-9. |  |
| Cheng, L., et al. (2015). "An evaluation of laboratory data at admission for predicting mortality among critically ill patients with cancer." *Cancer Research* 75(15). |  |
| Agulnik, A., et al. (2017). "Validation of a pediatric early warning system for hospitalized pediatric oncology patients in a resource-limited setting." *Cancer* 123(24): 4903-4913 |  |
| **Injury/surgical triage** | **N=3** |
| Chang, C. J., et al. (2013). "Predicting hospital mortality in adult patients with prolonged stay (>14 days) in surgical intensive care unit." *Minerva Anestesiol* 79(8): 843-852. |  |
| Kim, J., et al. (2009). "Development and validation of the Excess Mortality Ratio-adjusted Injury Severity Score Using the International Classification of Diseases 10th Edition." *Acad Emerg Med* 16(5): 454-464. |  |
| Muessig, J. M., et al. (2018). "Clinical Frailty Scale (CFS) reliably stratifies octogenarians in German ICUs: a multicentre prospective cohort study." *BMC Geriatr* 18(1): 162. |  |
| **Poor performing tool/score** | **N=5** |
| El Maraghi, S., et al. (2016). "Kallistatin level as a novel prognostic marker for community acquired pneumonia (CAP) in critically ill patients." *Intensive Care Medicine Experimental* 4. |  |
| Hsin, C. H., et al. (2016). "Venovenous extracorporeal membrane oxygenation in adult respiratory failure: Scores for mortality prediction." *Medicine (Baltimore)* 95(25): e3989. |  |
| Suberviola, B., et al. (2011). "Prognostic value of proadrenomedullin in severe sepsis and septic shock patients with community-acquired pneumonia." *Critical Care* 15: S98 |  |
| Suberviola, B., et al. (2013). "Hospital mortality prognostication in sepsis using the new biomarkers suPAR and proADM in a single determination on ICU admission." *Intensive Care Med* 39(11): 1945-1952. |  |
| Wong, H. R., et al. (2014). "A multibiomarker-based outcome risk stratification model for adult septic shock*." *Crit Care Med* 42(4): 781-789. |  |
| **Too small sample size** | **N=2** |
| Morton B, Nweze K, O'Connor J, Turton P, Joeke E, Blakey JD, et al. Oxygen exchange and C-reactive protein predict safe discharge in patients with H1N1 influenza. *Qjm.* 2017 Apr 1;110(4):227-32. |  |
| Tan L, et al. (2020) "Lymphopaenia predicts disease severity of COVID-19: a descriptive predictive study." *Signal Transduction and Targeted Therapy* 5:33 |  |
| **Reviews – Reference list checked** | **N=1** |
| Wynants L, et al., (2020) "Prediction models for diagnosis and prognosis of covid-19: systematic review and critical appraisal". *BMJ* 369: m1328. |  |
| **Not in English** | **N=1** |
| Piccini, M., et al. (2010) Ethical, deontologic and legal considerations about SIAARTI Document "Clinical ethics recommendations for the allocation of intensive care treatments, in exceptional, resource-limited circumstances" |  |

**Supplement 1, Table S1.3.** Characteristics of publications informing ICU Triage <u>in pandemic</u> situations. N= 16.

| Author and publication year | Country of study subjects or where guideline applied | Publication type |  |  | Target population |  |  | Pathological Process |  | Proposed Benefit criteria |  |  |
| --- | --- | --- | --- | --- | --- | --- | --- | --- | --- | --- | --- | --- |
|  |  | Guideline/ consensus | Retro/prospective | Literature Review | Age range (years)<sup>a</sup> | Admitted to ICU | ED/other hospital | Pneumonia/Lung | Multi-organ/sepsis | ICU Admission/ not | Discharge (ICU) | Ventilator access |
| CONSENSUS |  |  |  |  |  |  |  |  |  |  |  |  |
| Christian et al., 2006 | Canada | ✓ |  |  | 0-paediatric | ✓ | ✓ | ✓ |  | ✓ | ✓ |  |
| Devereaux et al, 2007 | USA | ✓ |  |  | All ages | ✓ |  |  |  | ✓ | ✓ |  |
| Powell et al., 2008 | USA | ✓ |  |  | 0 - older adult | ✓ | ✓ | ✓ | ✓ | ✓ | ✓ | ✓ |
| White et al., 2009 | USA | ✓ |  |  | 12- older adult |  | ✓ | ✓ | ✓ |  |  | ✓ |
| Wilkens et al., 2010 | USA | ✓ |  |  | 0- older adult |  | ✓ | ✓ | ✓ |  |  | ✓ |
| NewYork Taskforce, 2015 | USA | ✓ |  |  | 0- older adult | ✓ |  | ✓ | ✓ | ✓ | ✓ | ✓ |
| Chuang et al., 2020 | USA | ✓ |  |  | Adults | ✓ |  |  | ✓ | ✓ | ✓ | ✓ |
| ALGORITHMS/FACTORS |  |  |  |  |  |  |  |  |  |  |  |  |
| Guest et al, 2009 | UK |  | P |  | 60 (IQR 41-69) | ✓ |  | ✓ |  | ✓ |  |  |
| Khan et al., 2009 | UK |  | R |  | 26-52 | ✓ |  |  | ✓ | ✓ | ✓ |  |
| Grissom et al., 2010 | USA |  | R |  | 50 ±20 | ✓ |  |  | ✓ |  | ✓ | ✓ |
|  |  |  | P |  | 53 ±20 |  |  |  |  |  |  |  |
| Chuang et al, 2020 | USA |  | P |  | All ages | ✓ |  | ✓ |  | ✓ |  | ✓ |
| Adenijil et al., 2011 | UK |  | R |  | 18-71 | ✓ |  | ✓ |  | ✓ |  |  |
| Cummings et al., 2020 | USA |  | P |  | 62 (51–72) | ✓ |  | ✓ | ✓ |  |  |  |
|  |  | Guideline/ consensus | Retro/prospective | Literature Review | Age range (years)<sup>α</sup> | Admitted to ICU | ED/other hospital | Pneumonia/Lung | Multi-organ/sepsis | ICU Admission/ not | Discharge (ICU) | Ventilator access |
| Talmor et al., 2007 | USA |  | R |  | Mean 59 (≥ 18) |  | ✓ |  | ✓ | ✓ |  | ✓ |
| Liang et al., 2020 | China |  | R |  | 48.2±15.2 (4-88) |  | ✓ | ✓ |  | ✓ |  |  |
| Semple et.al., 2013 | UK |  | R |  | 0-older adult |  | ✓ | ✓ | ✓ |  |  | ✓ |
| Kim et.al., 2012 | USA |  |  | LR | 0-paediatric | ✓ | ✓ | ✓ | ✓ | ✓ | ✓ | ✓ |

<sup>α</sup> range or mean ±SD or median and IQR as reported in eligibility or findings sections of eligible articles
(D): Derivation cohort; DK=Denmark; LR=Literature review; N/R= Not reported ; SOFA= Sepsis-related Organ Failure Assessment P= Prospective; R=Retrospective;

**Supplement 1, Table S1.4.**
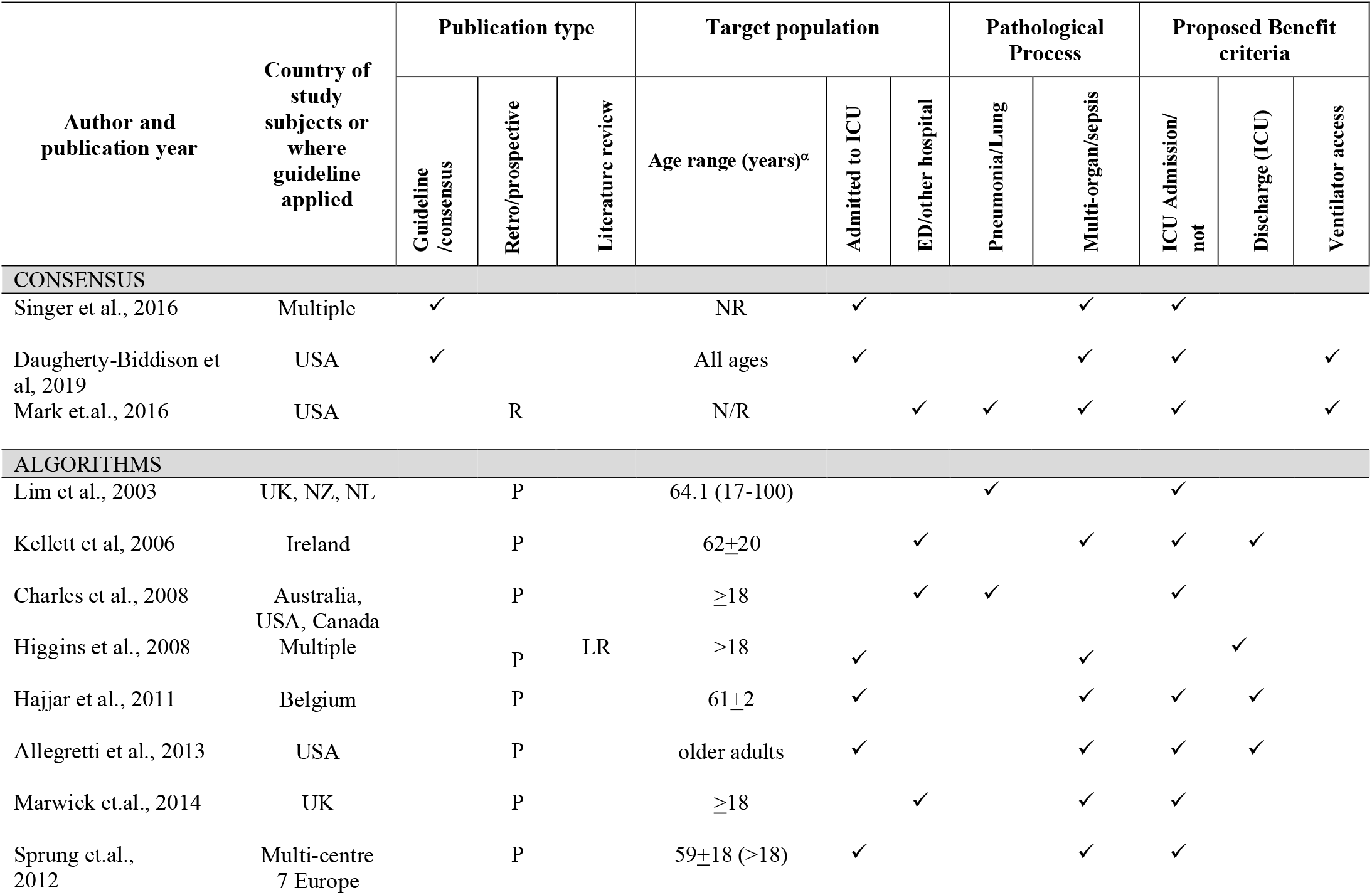
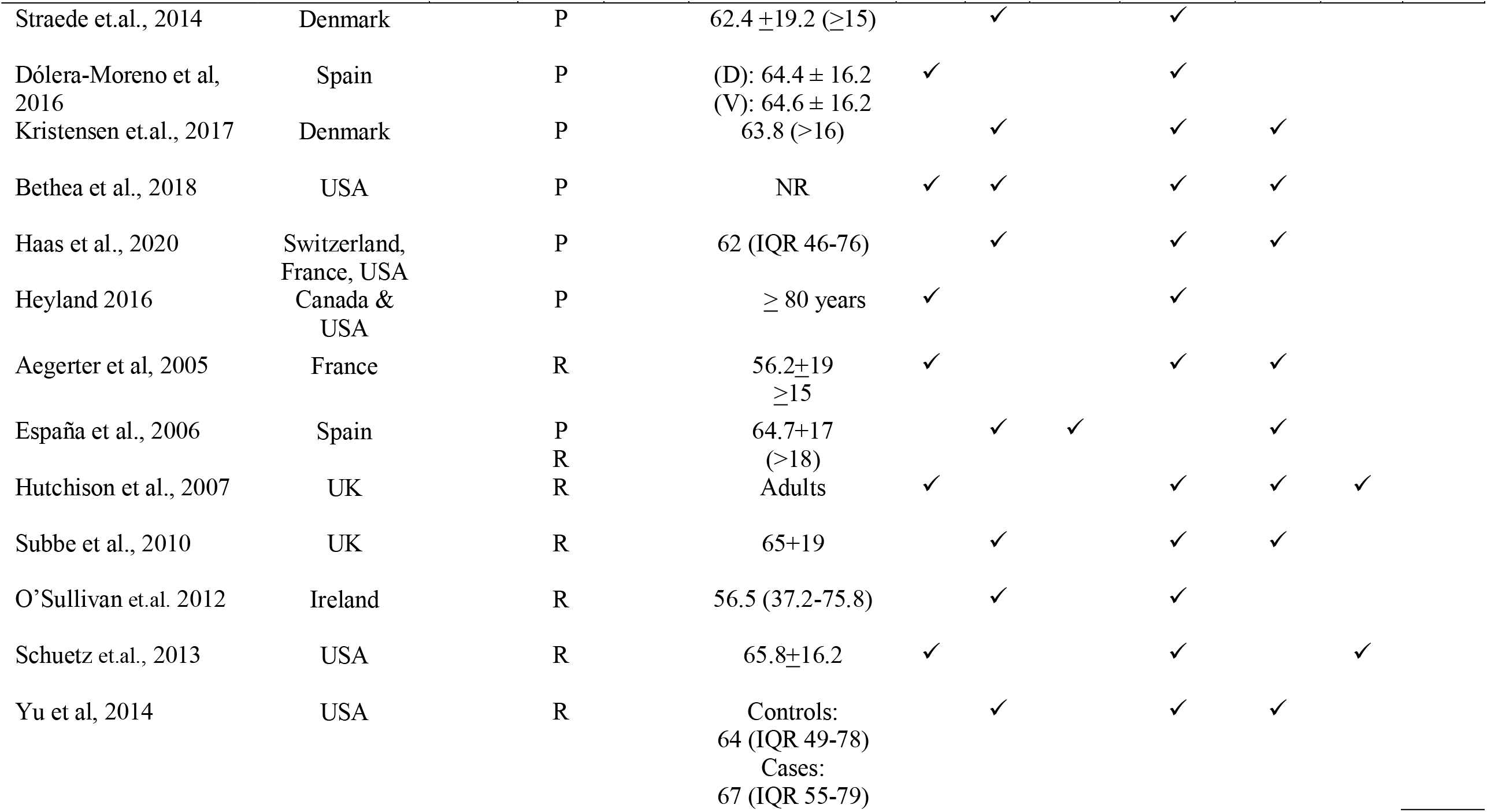
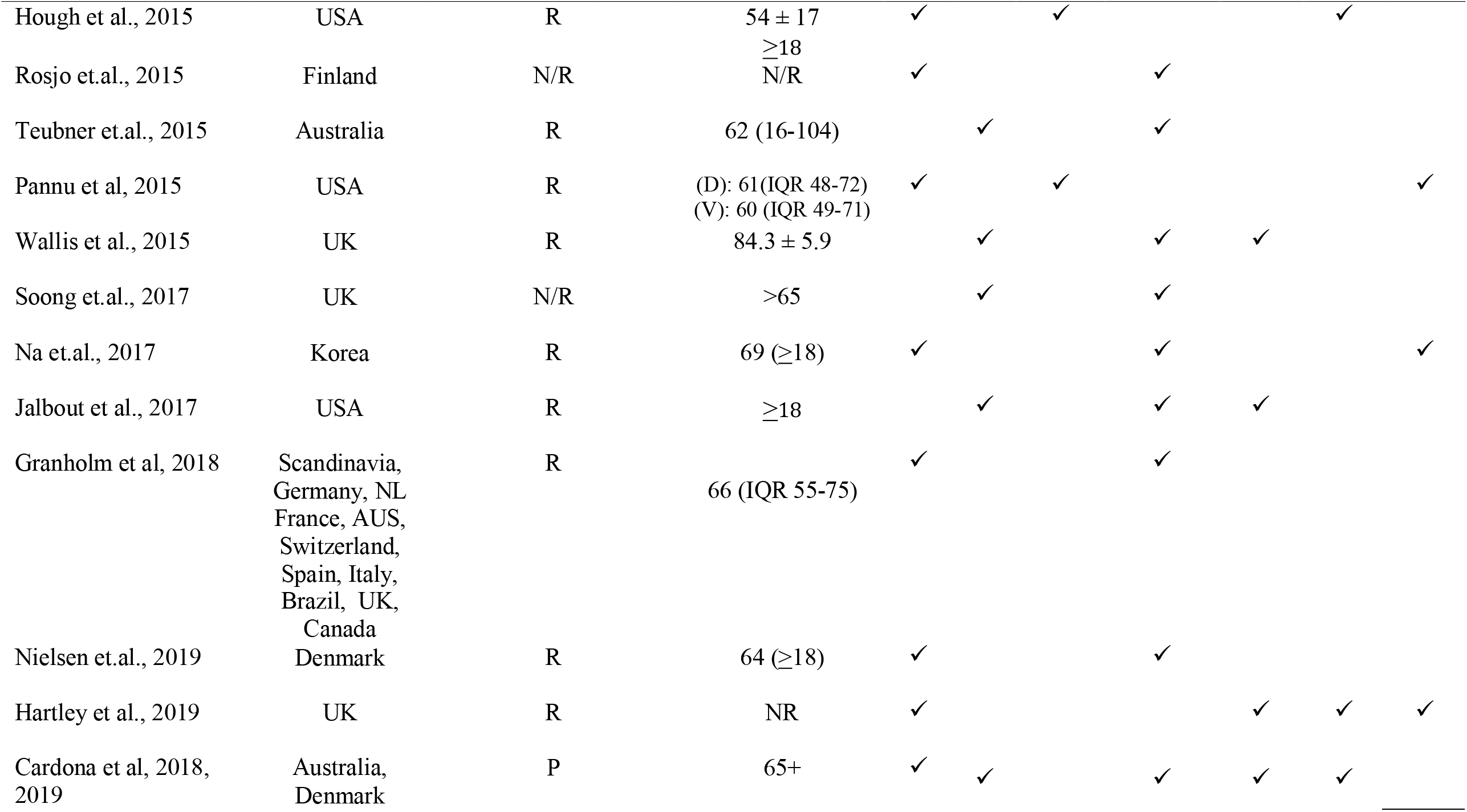
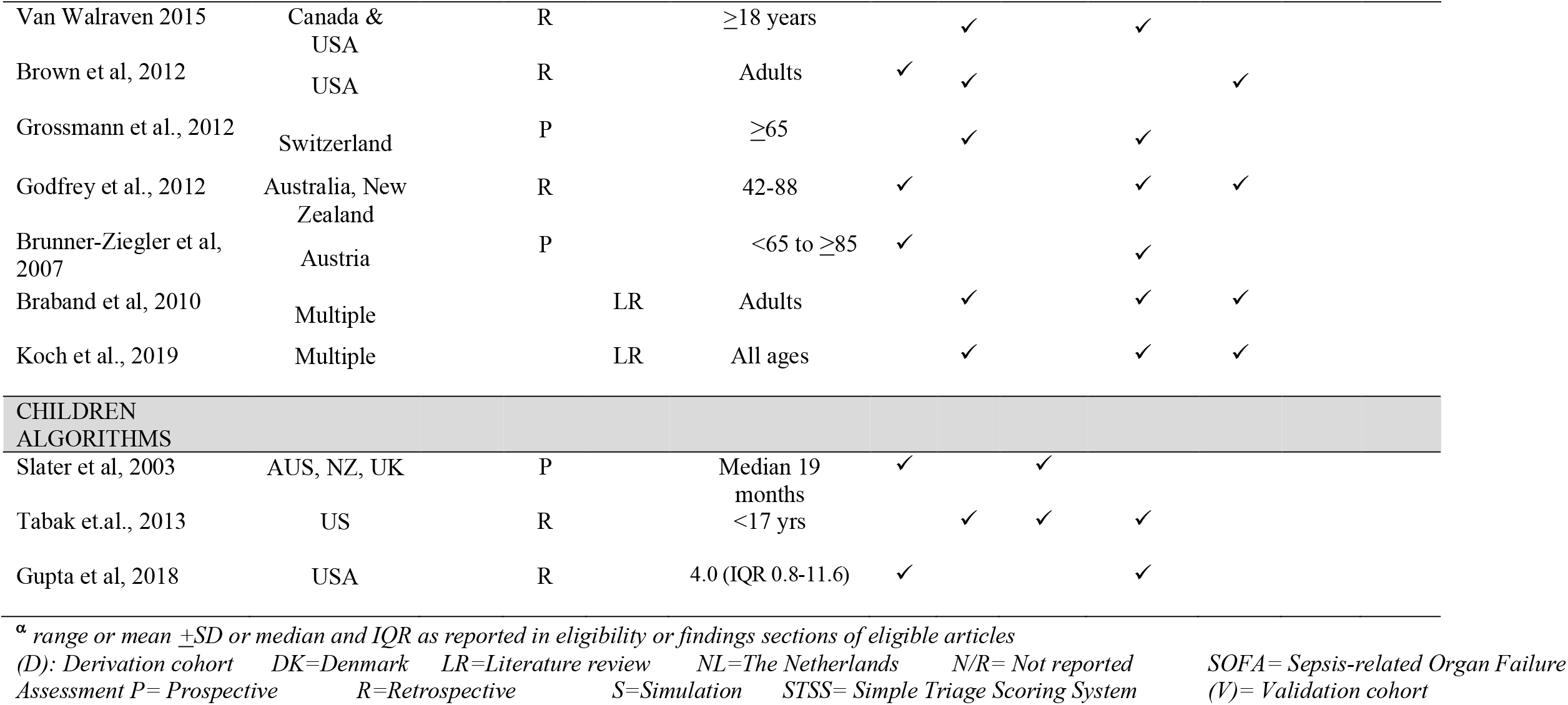
**Characteristics of publications on prognostic tools for risk of death in routine clinical care potentially applicable to ICU triage in pandemics (N= 46)**

**Supplement 2. Table S2.1.**
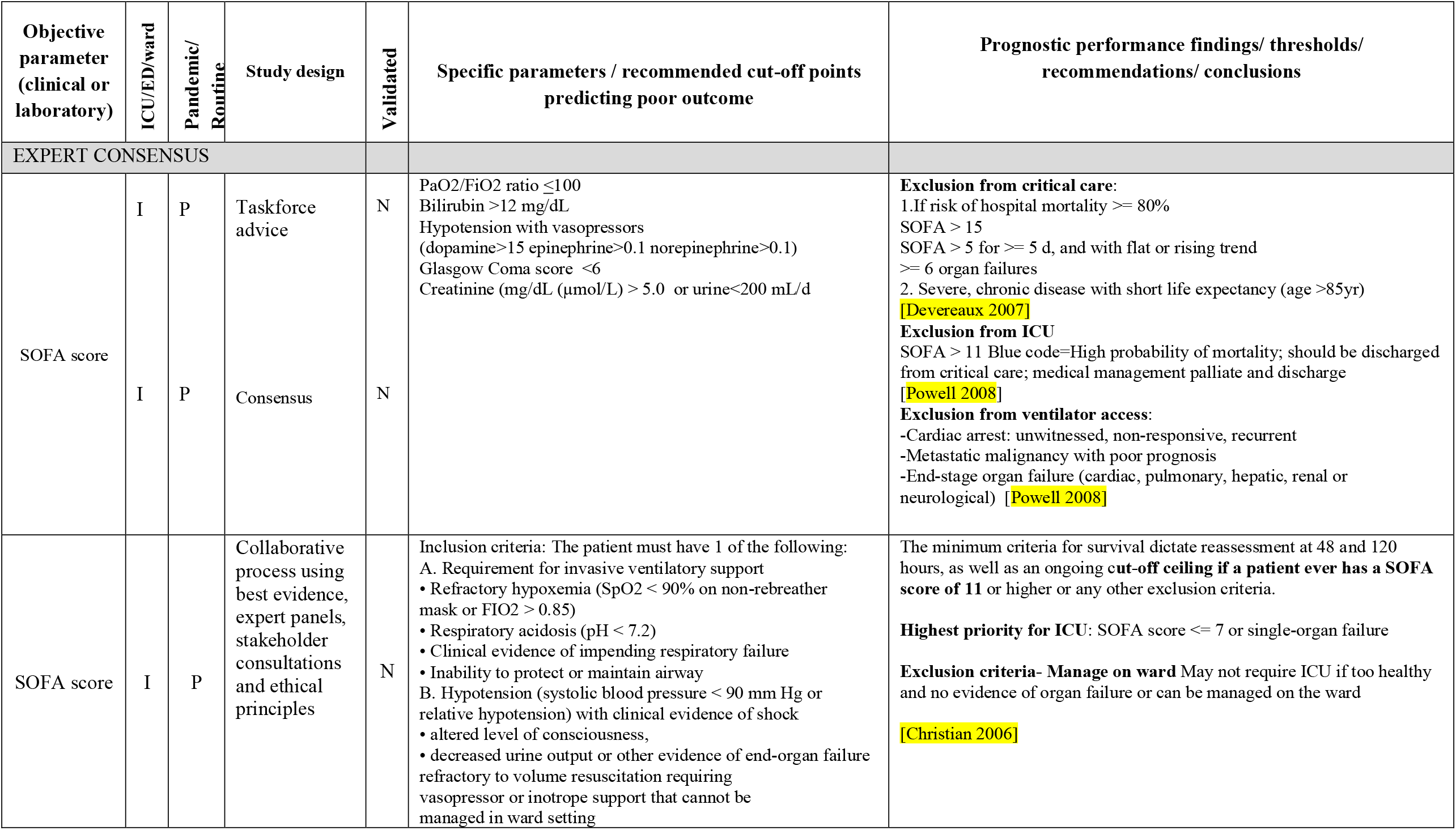
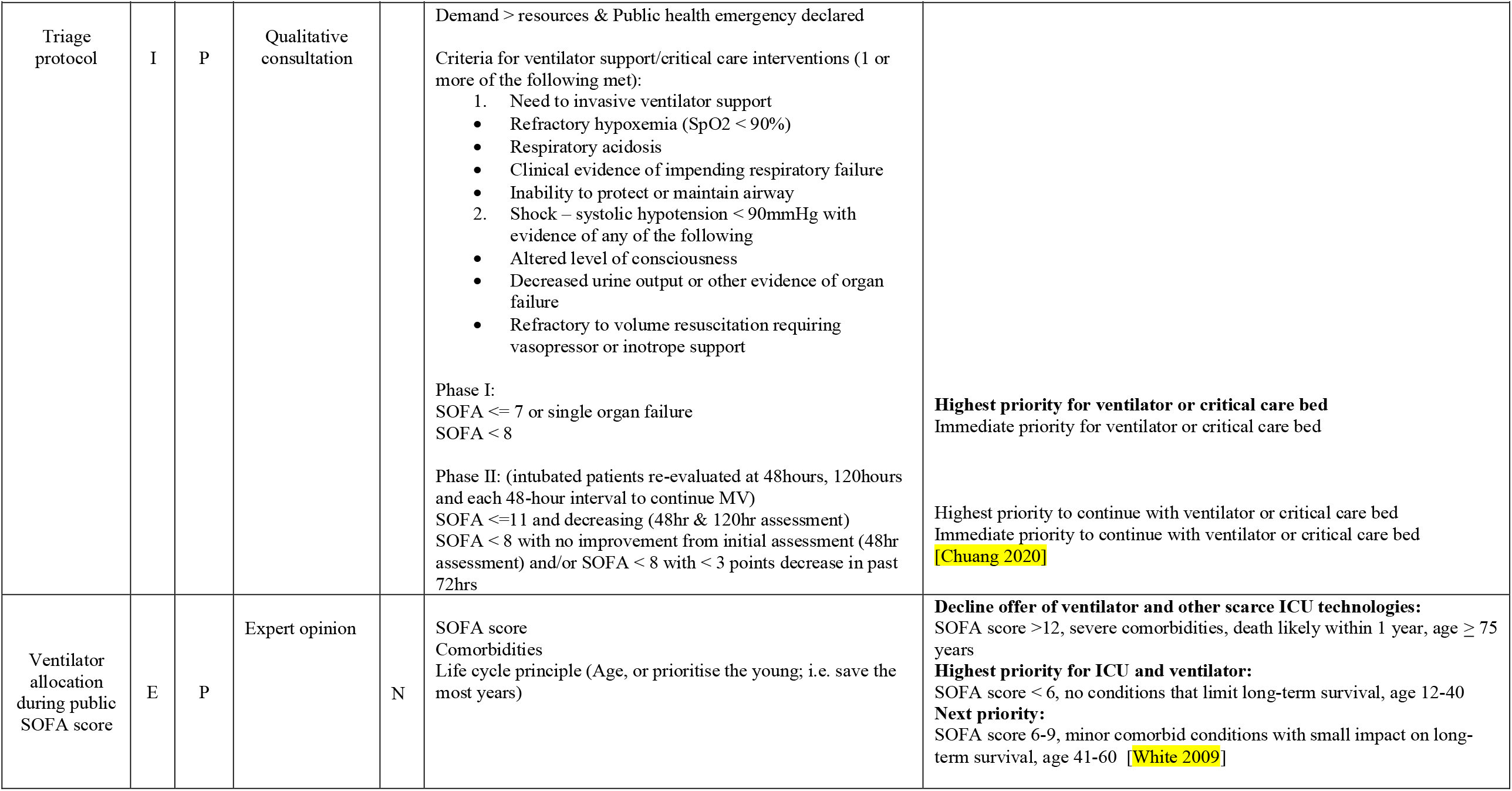
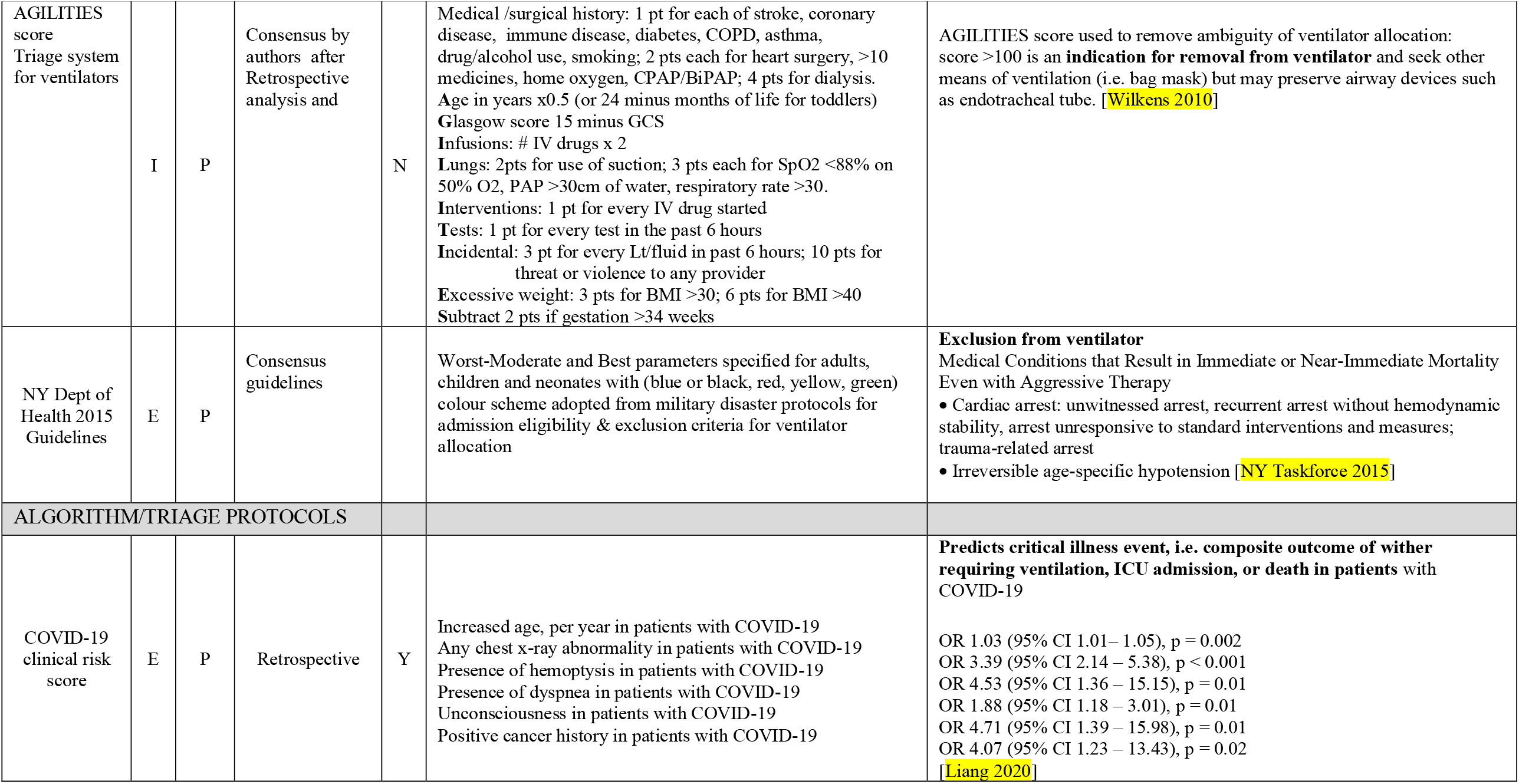
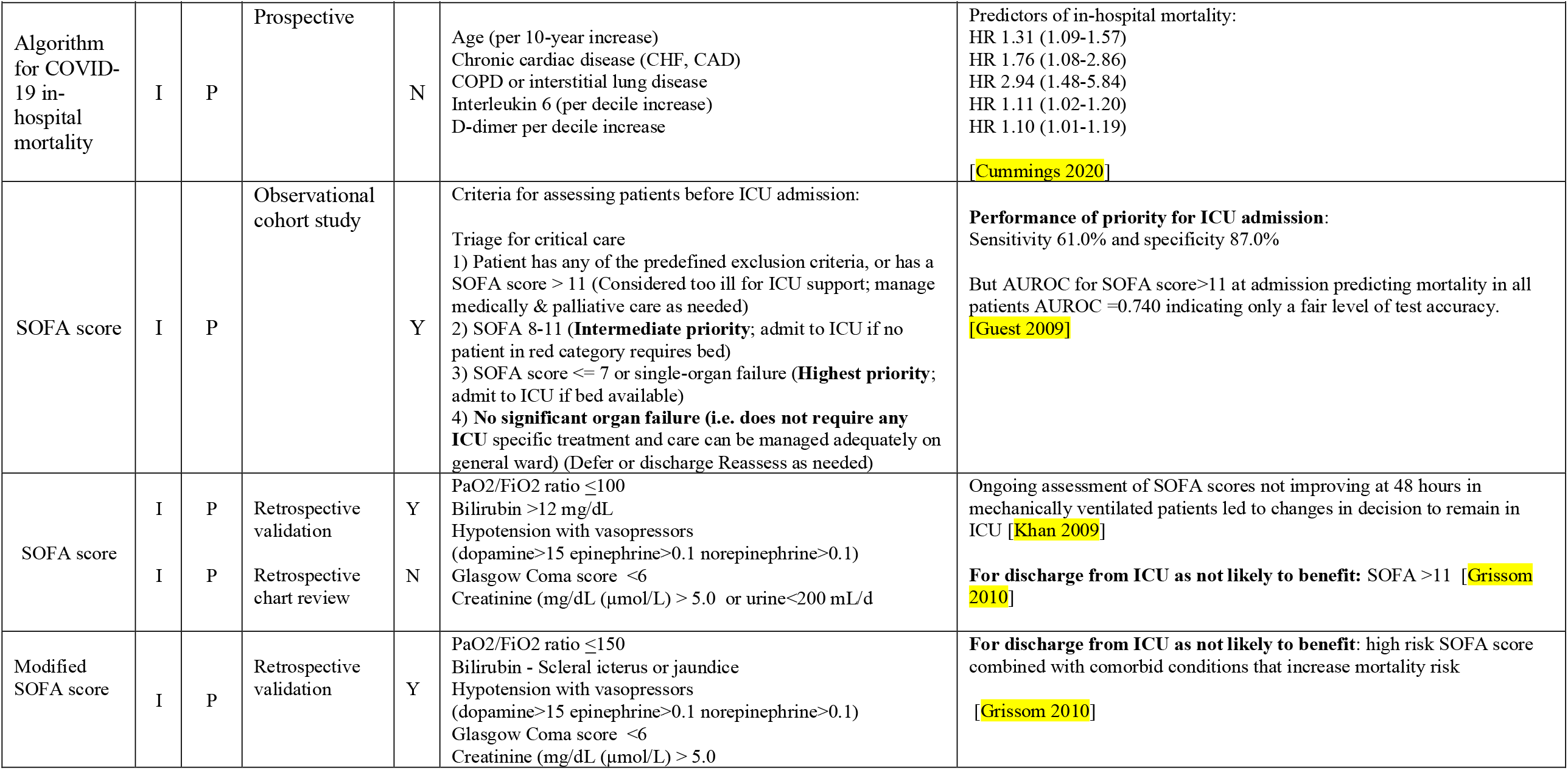
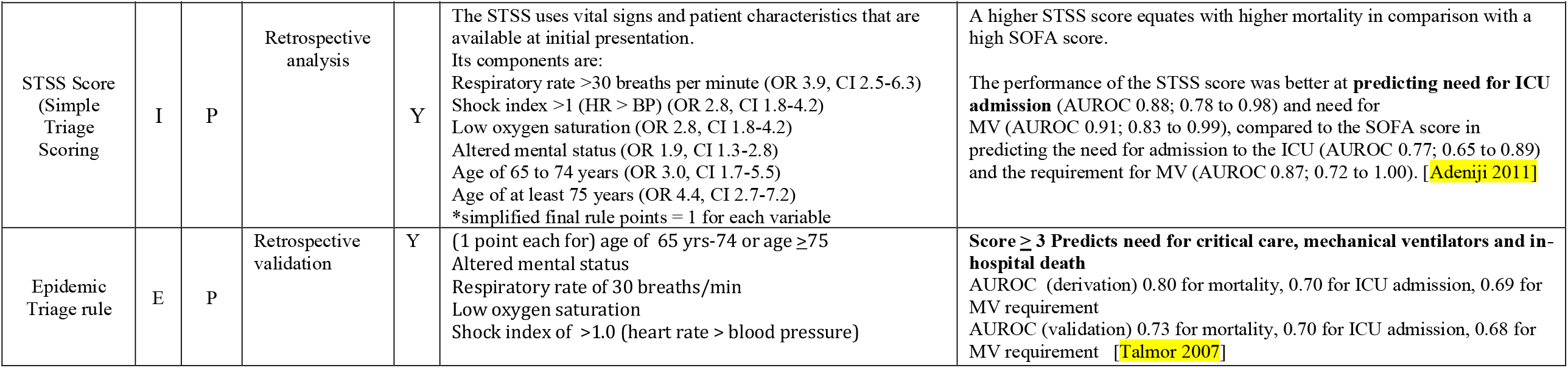
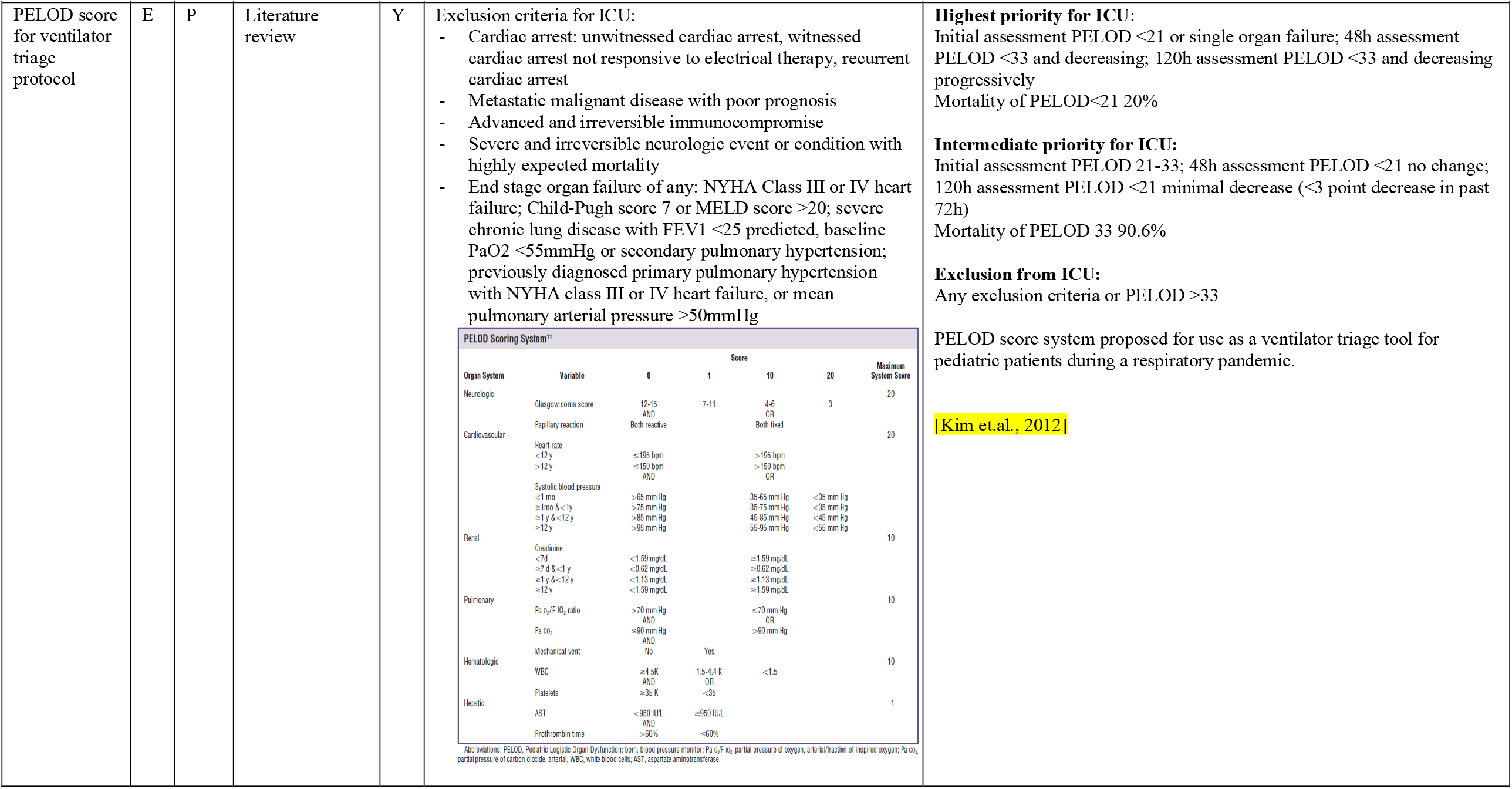
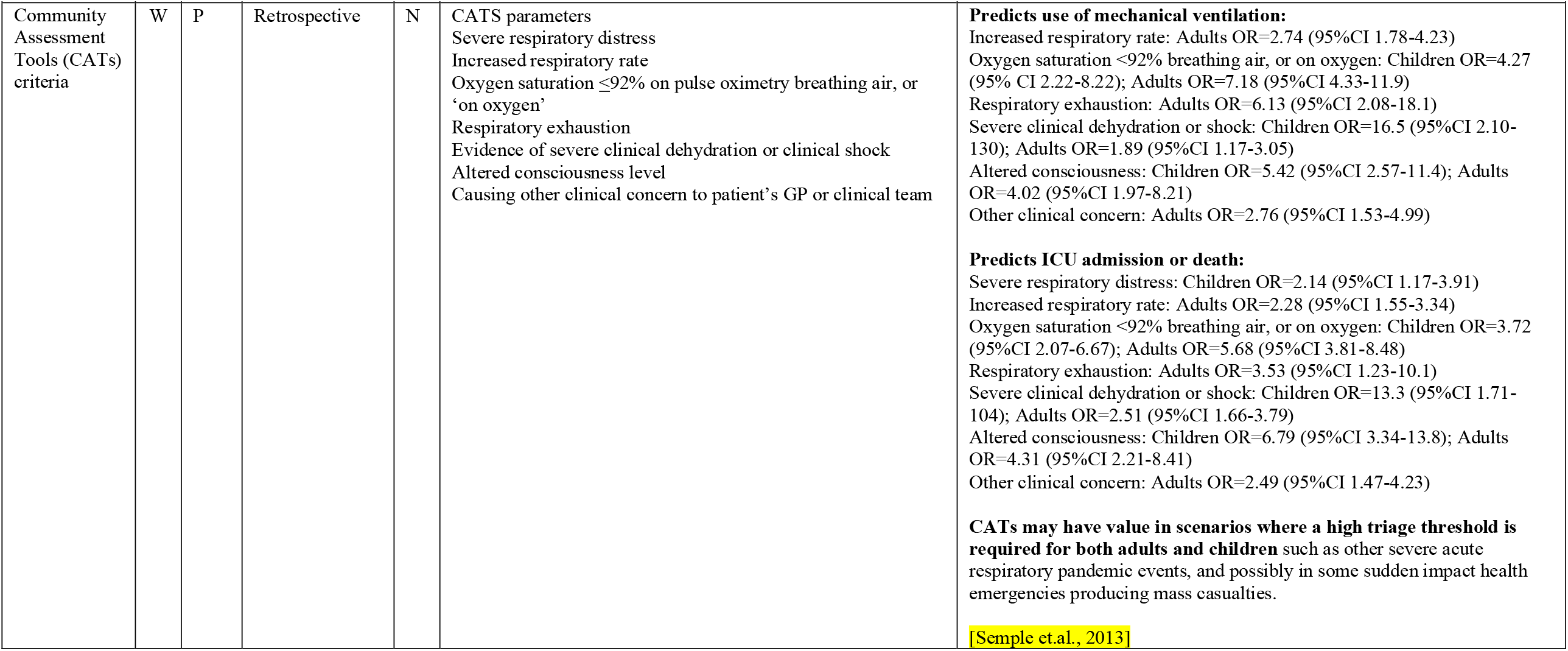
**Validation status, study types and clinical parameters predicting poor outcome for potentially guidance on ICU admission or discharge during pandemics (N=16)**

**Supplement 2. Table S2.2.**
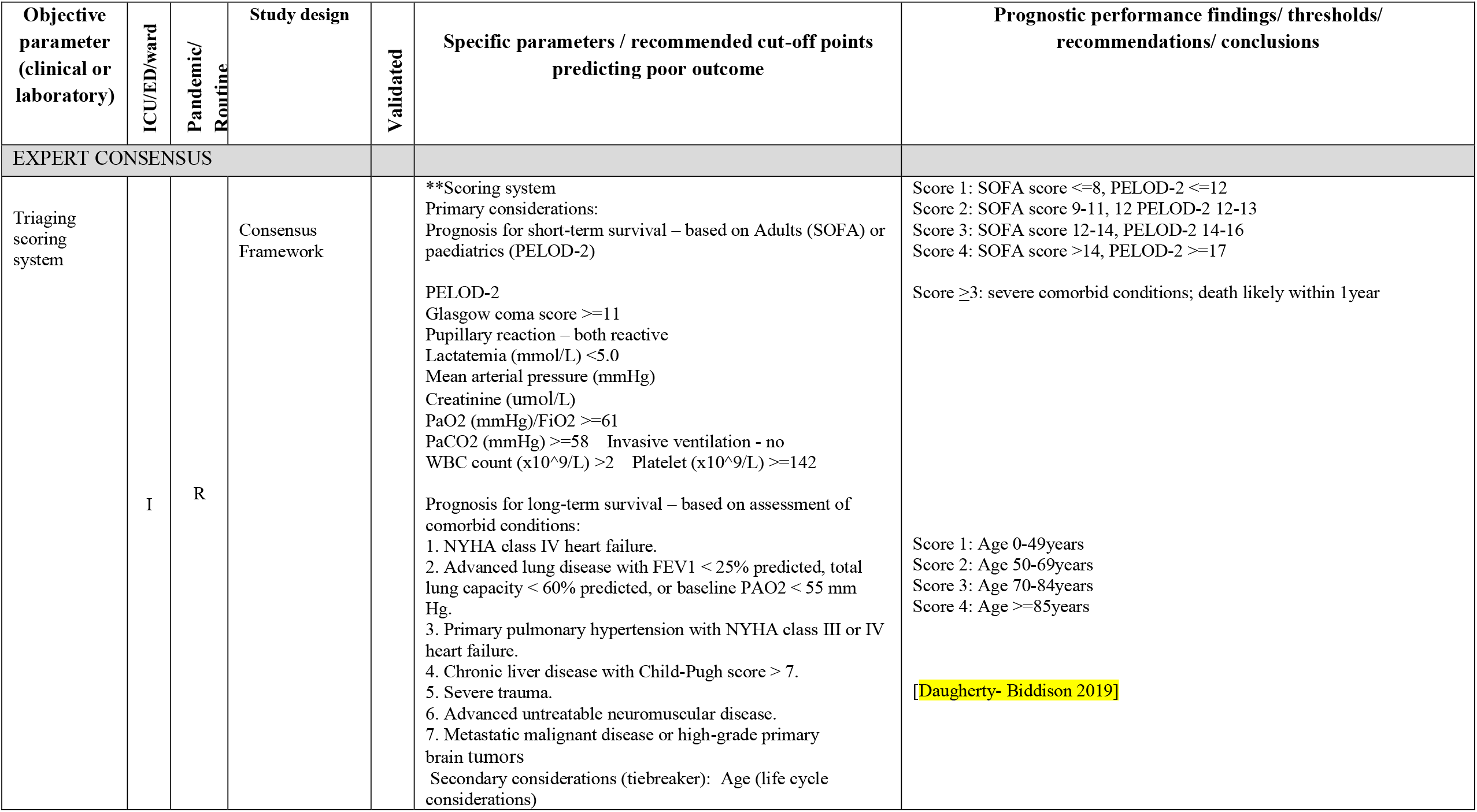
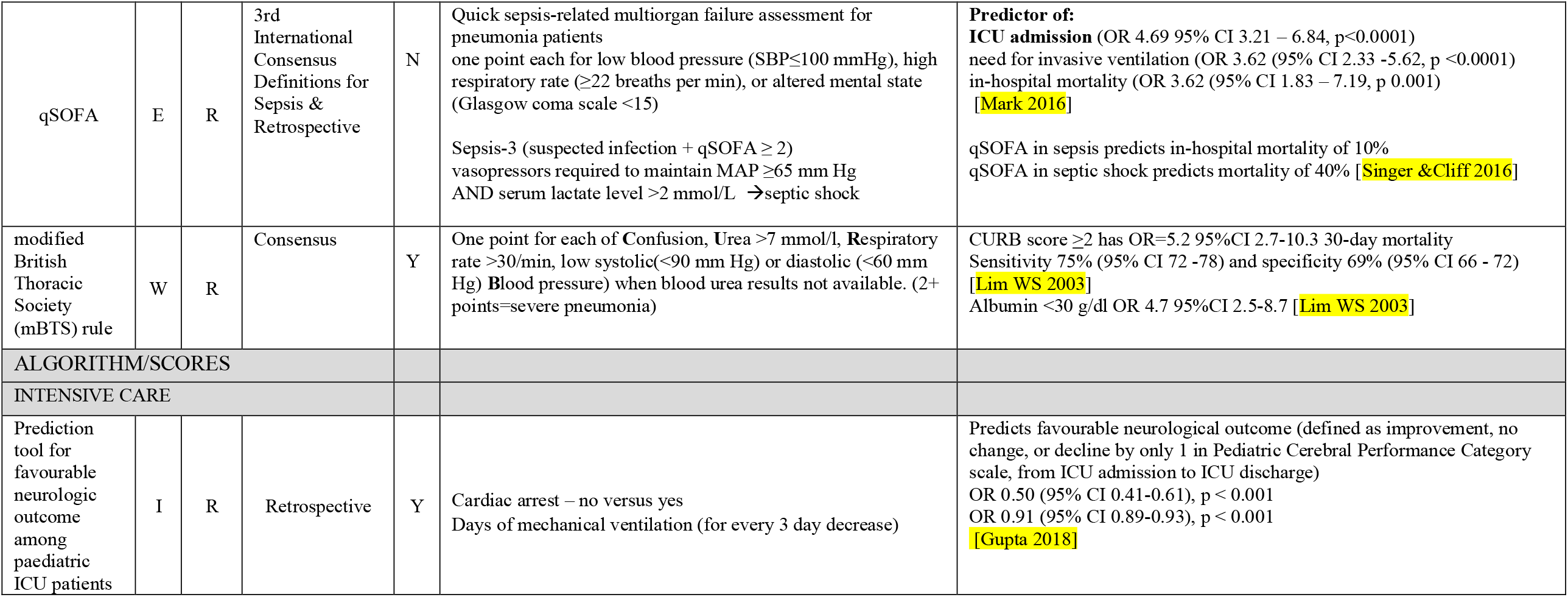
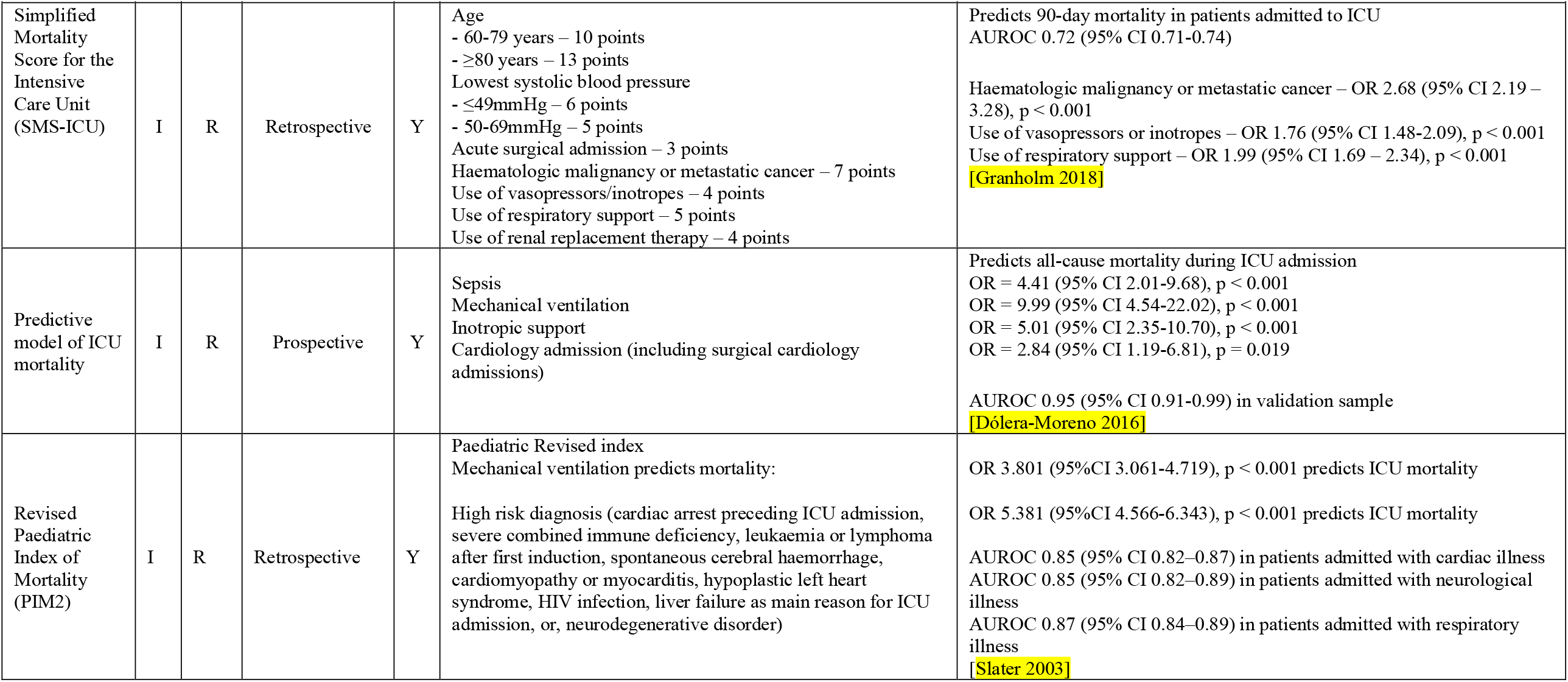
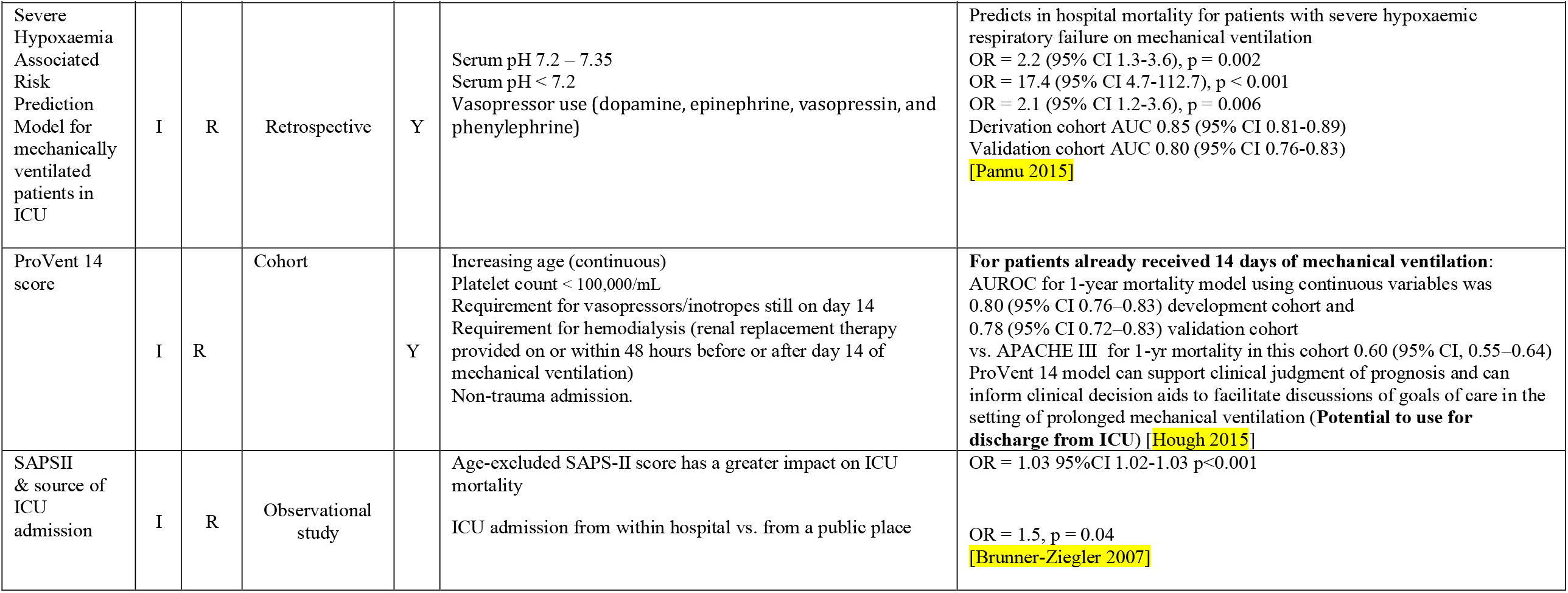
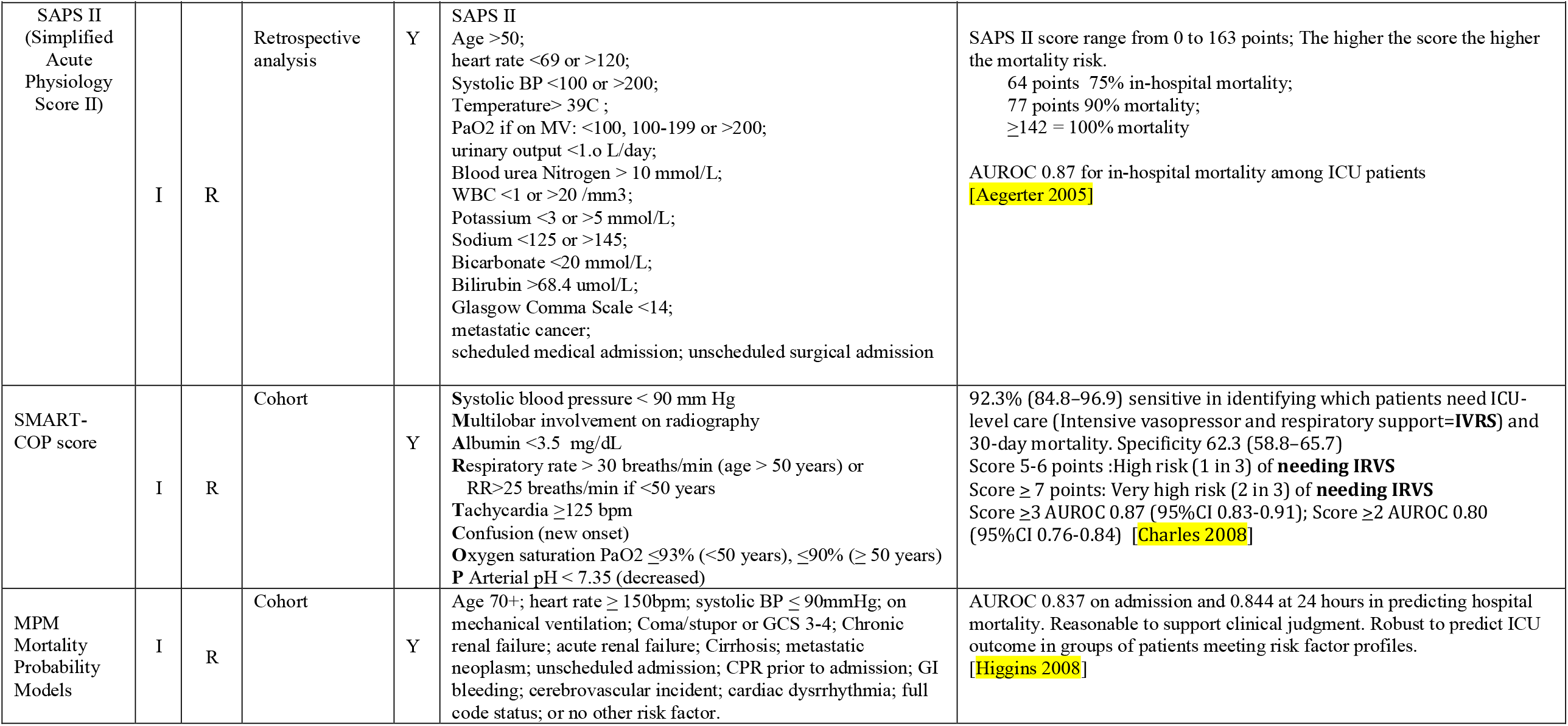
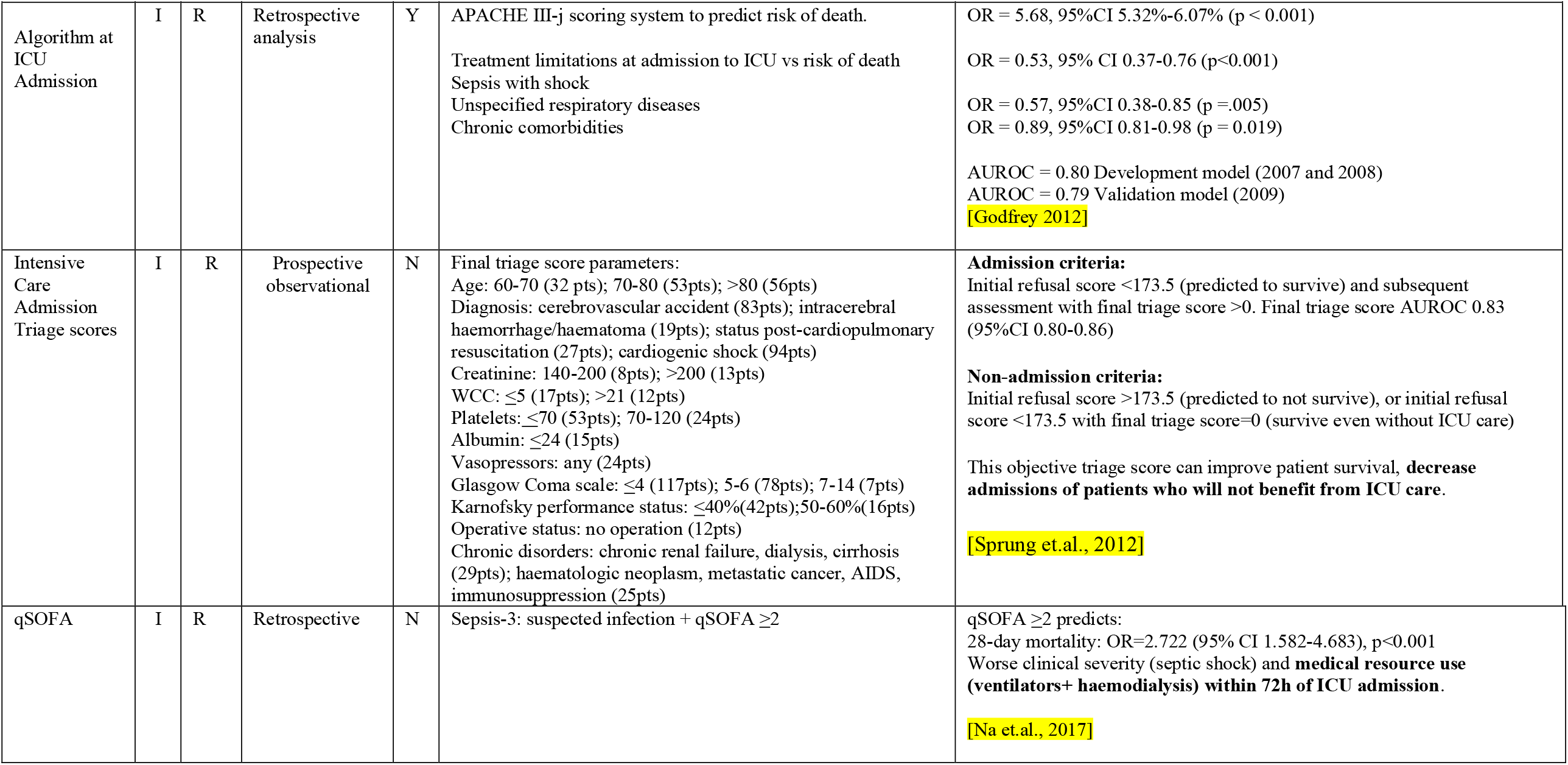
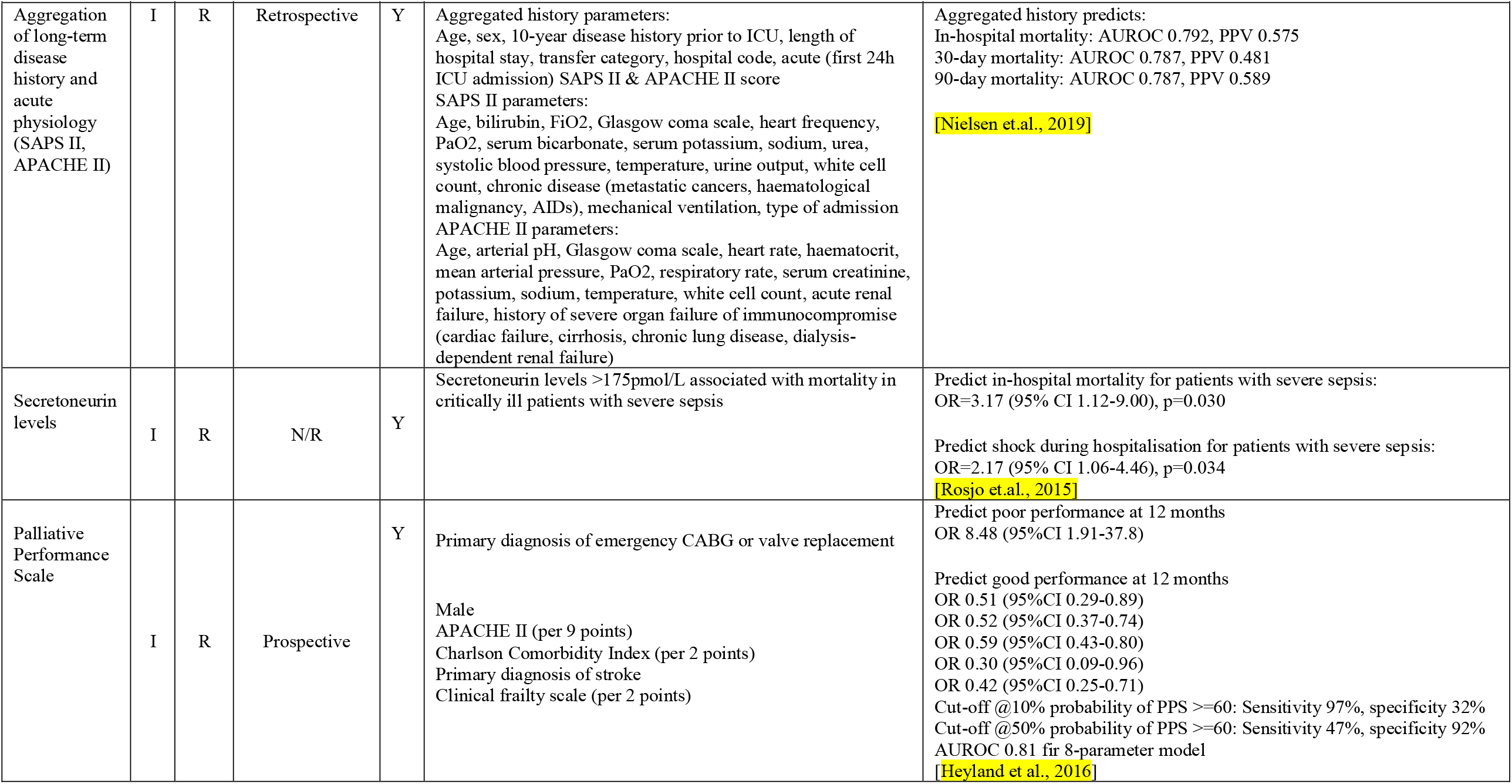
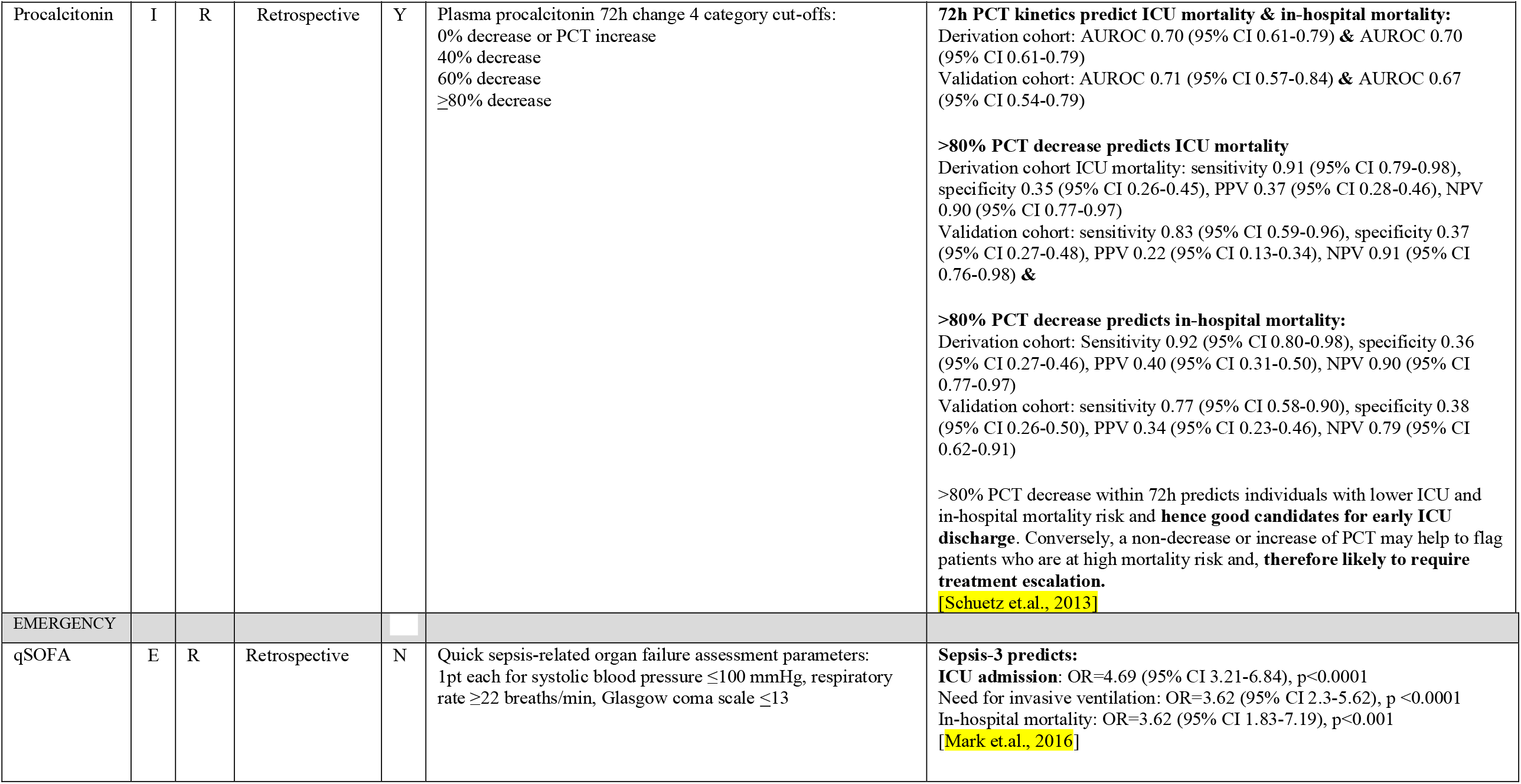
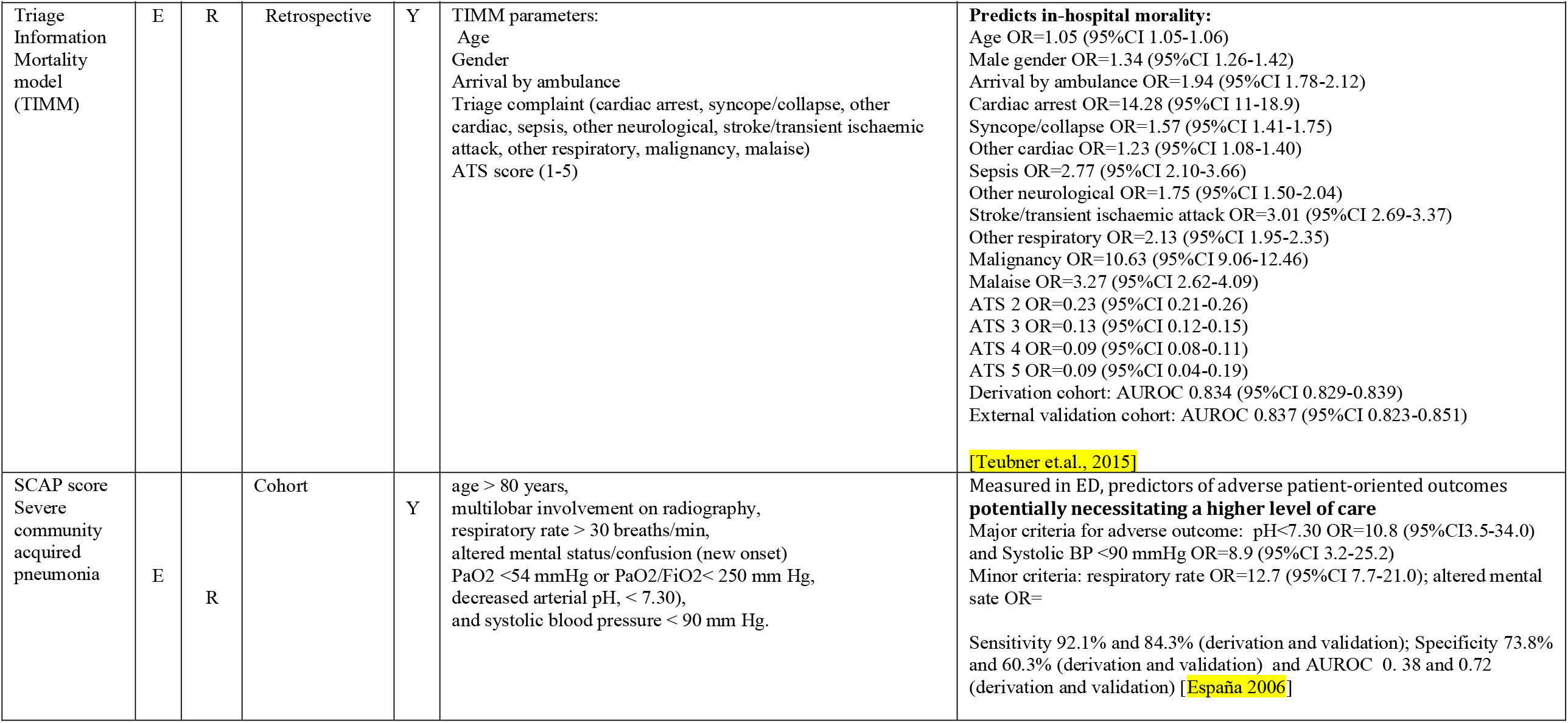
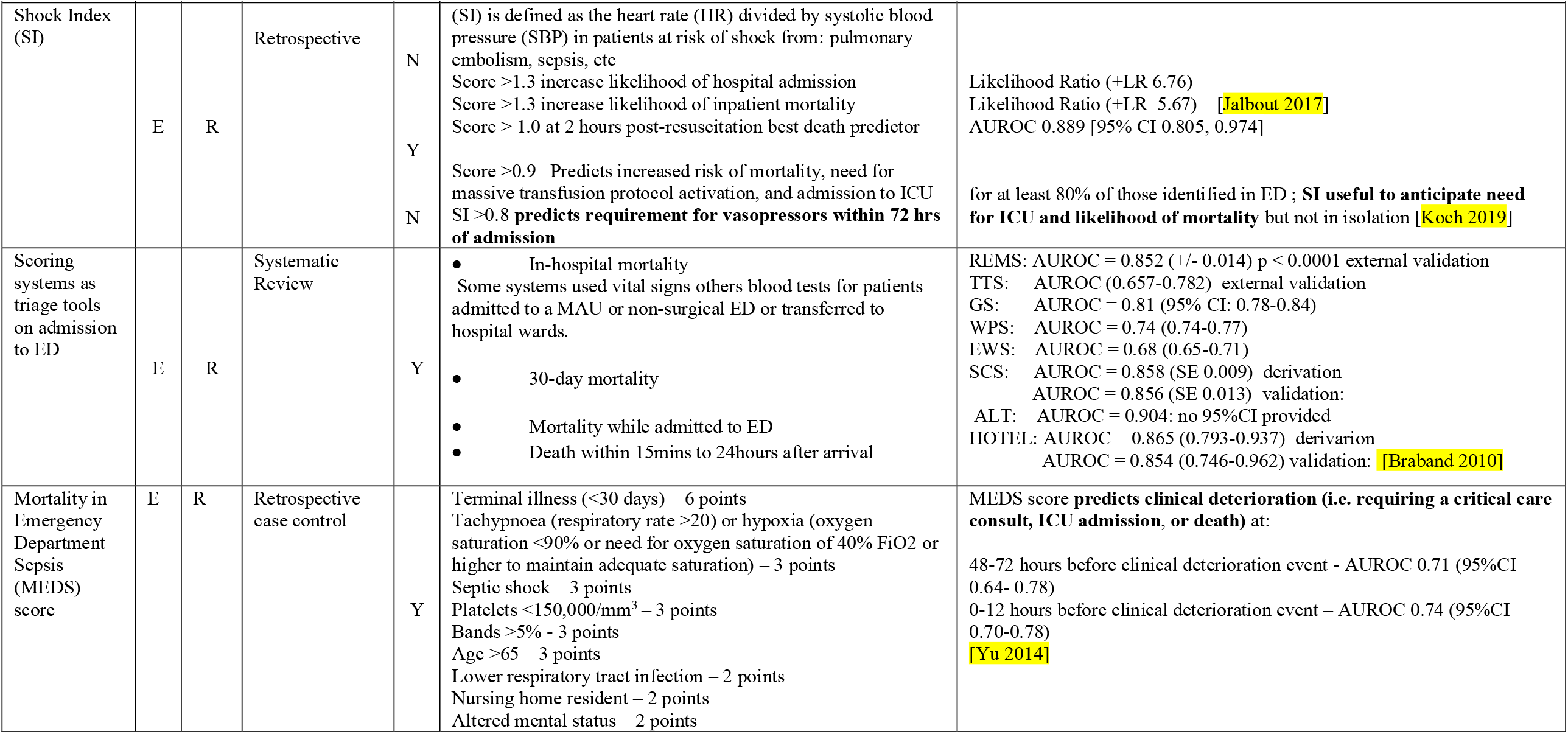
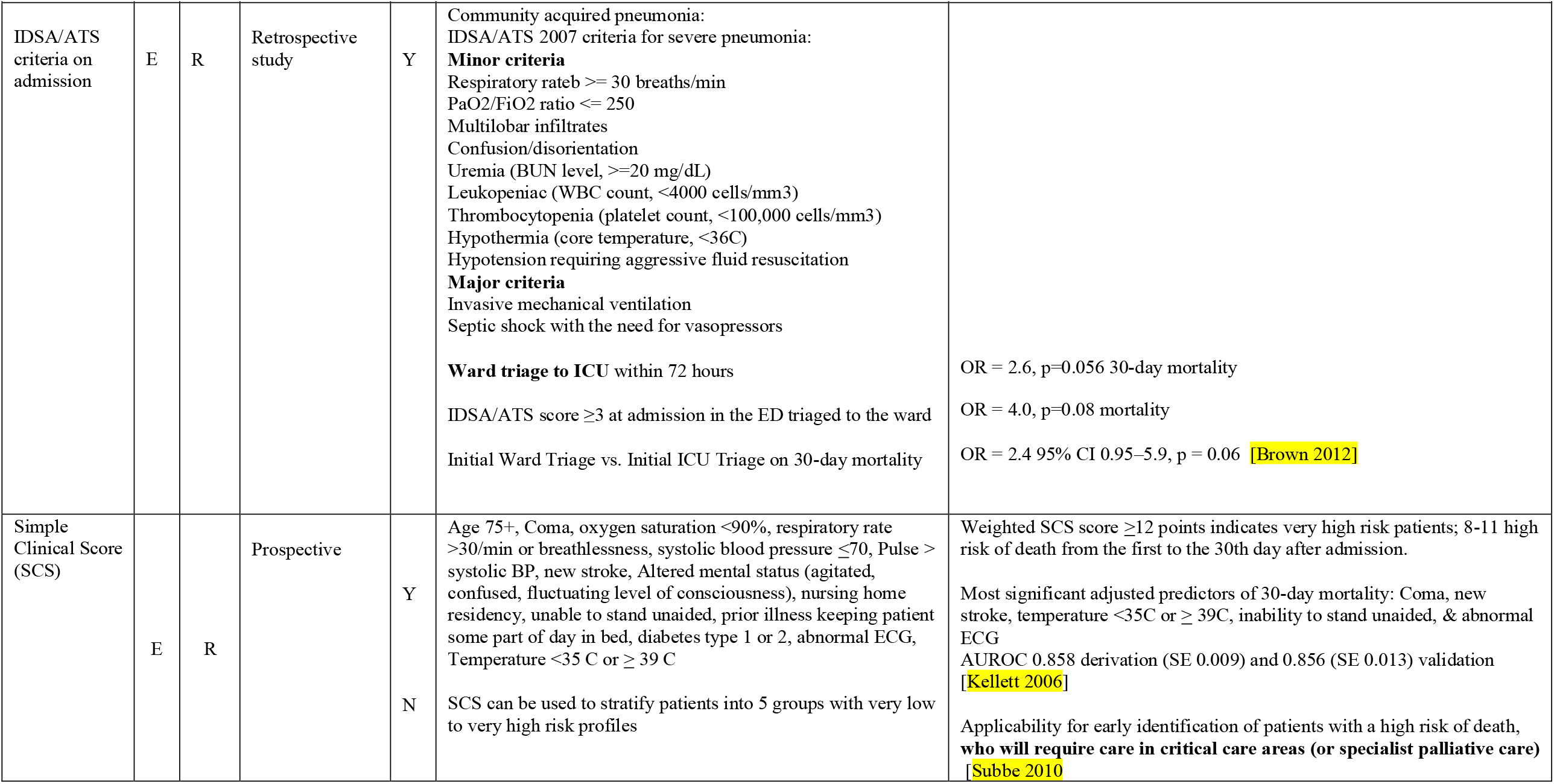
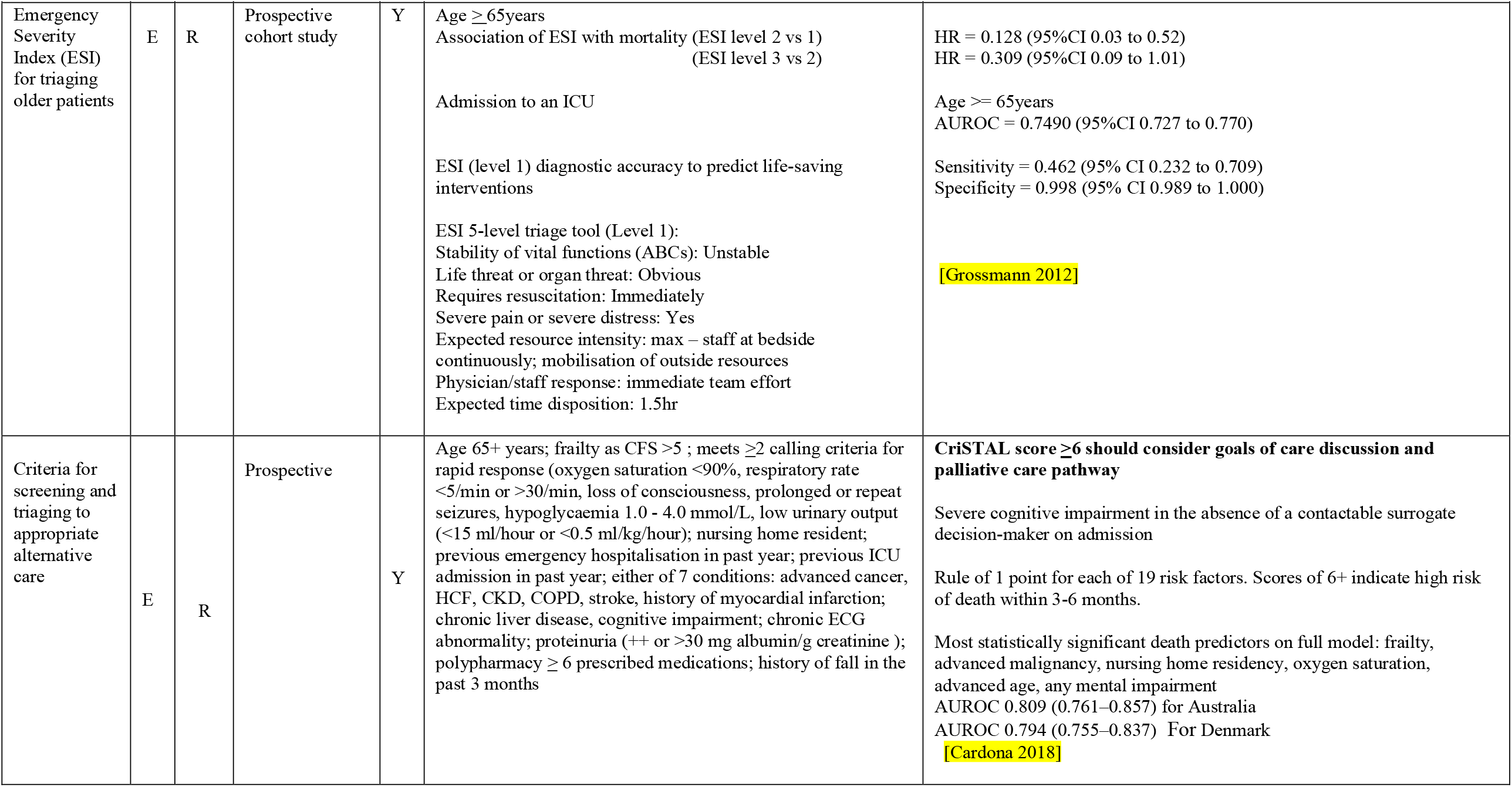
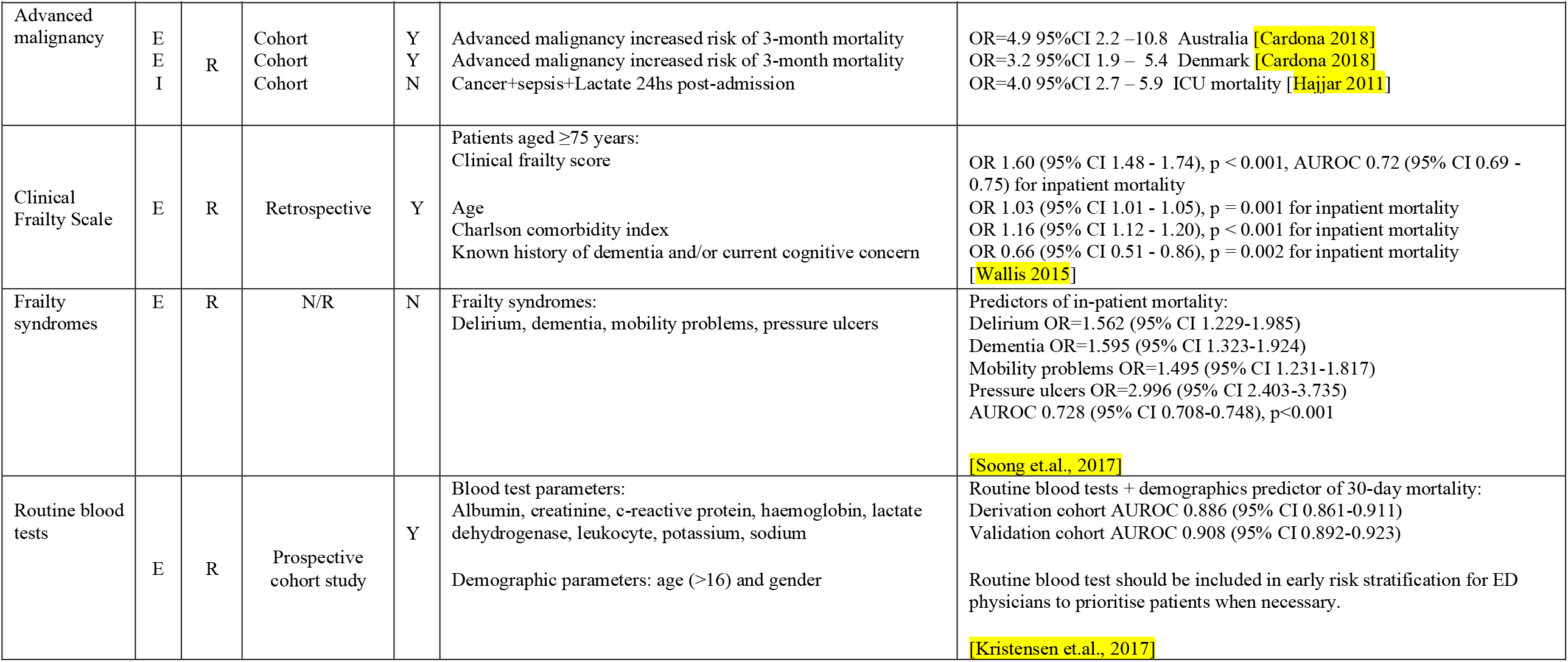
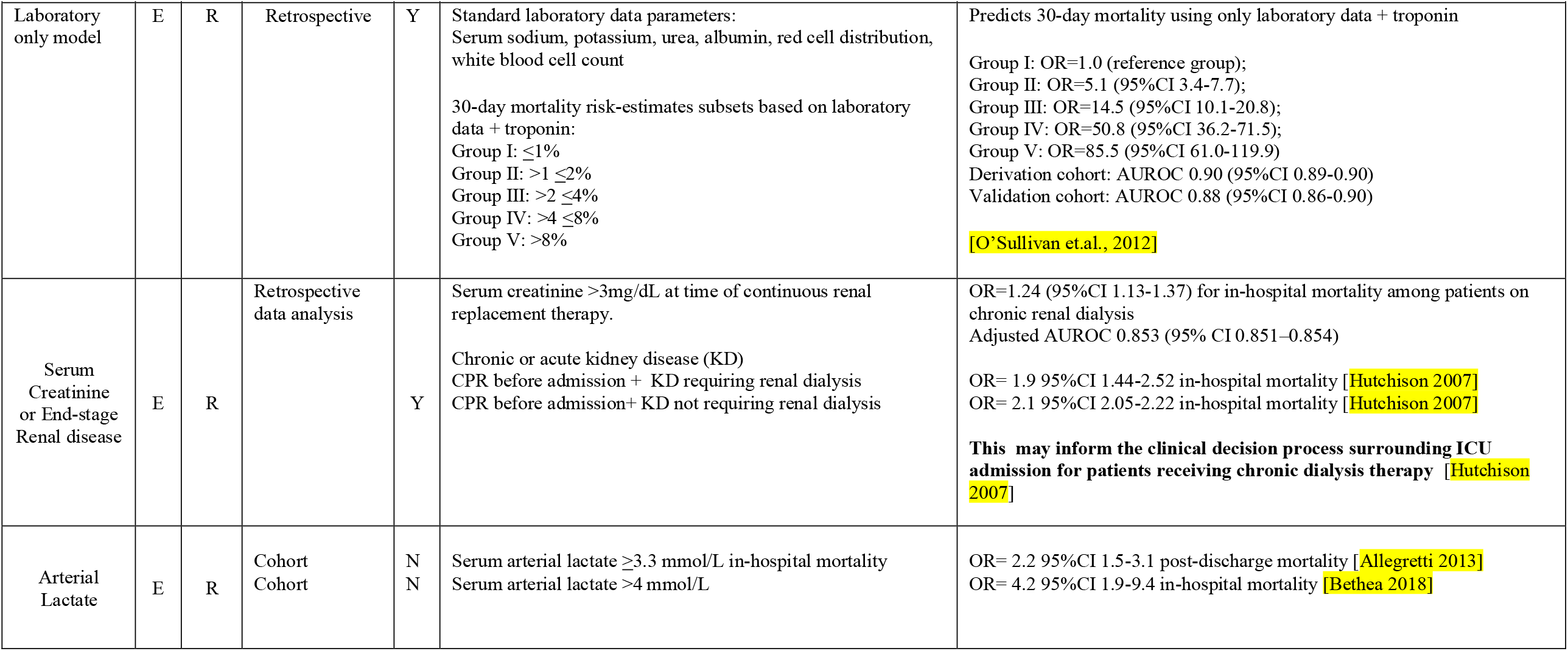
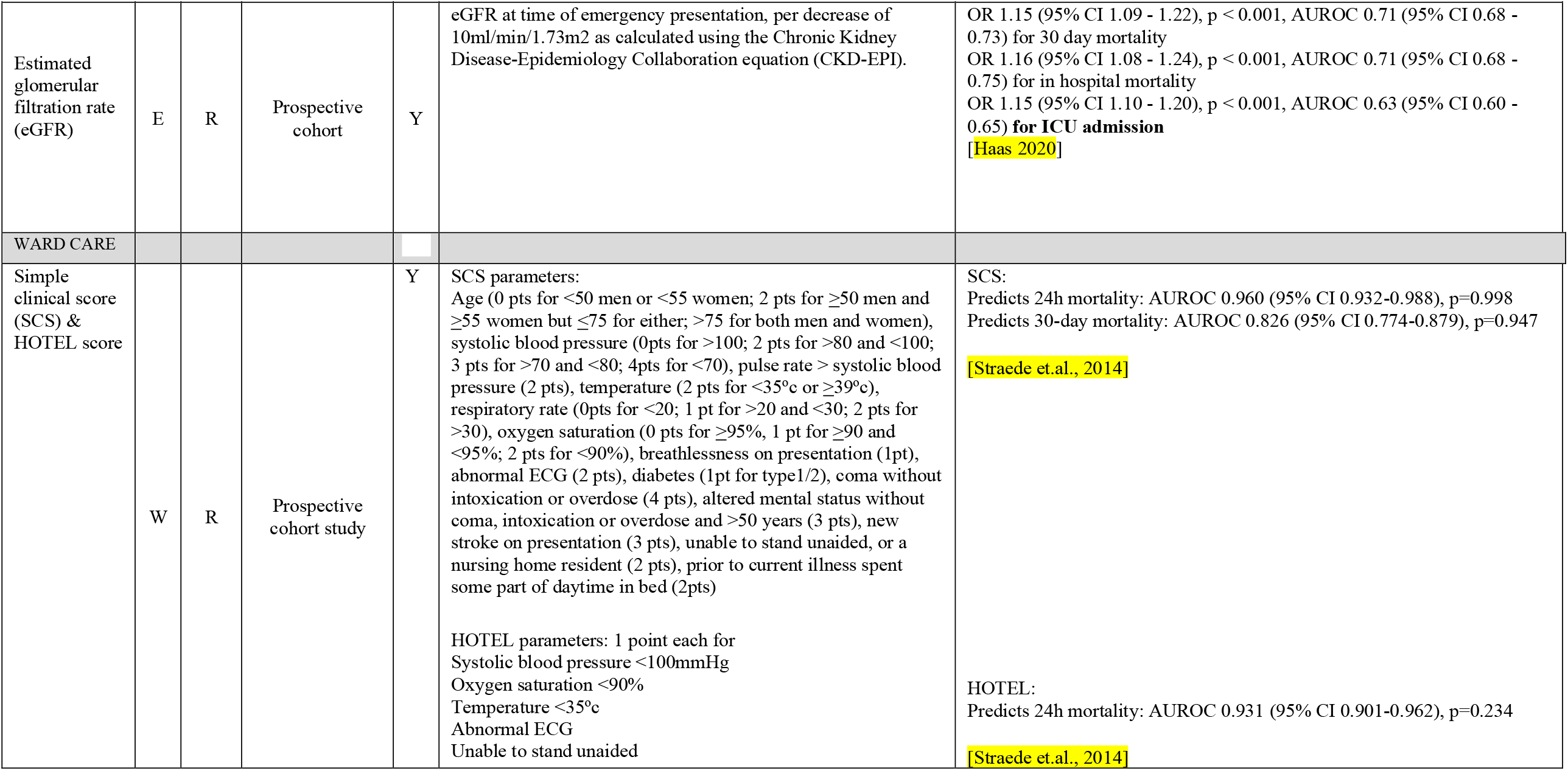
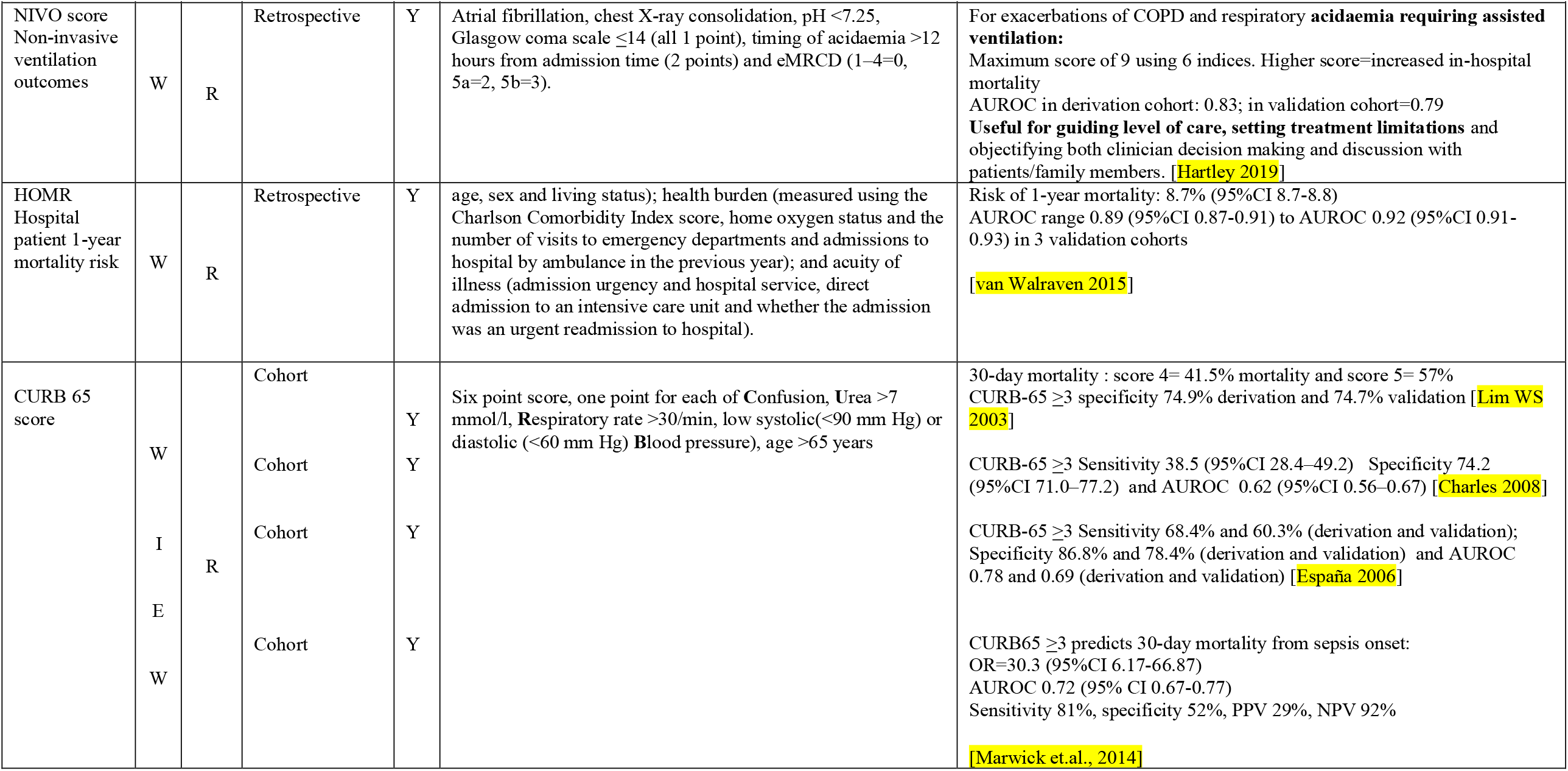
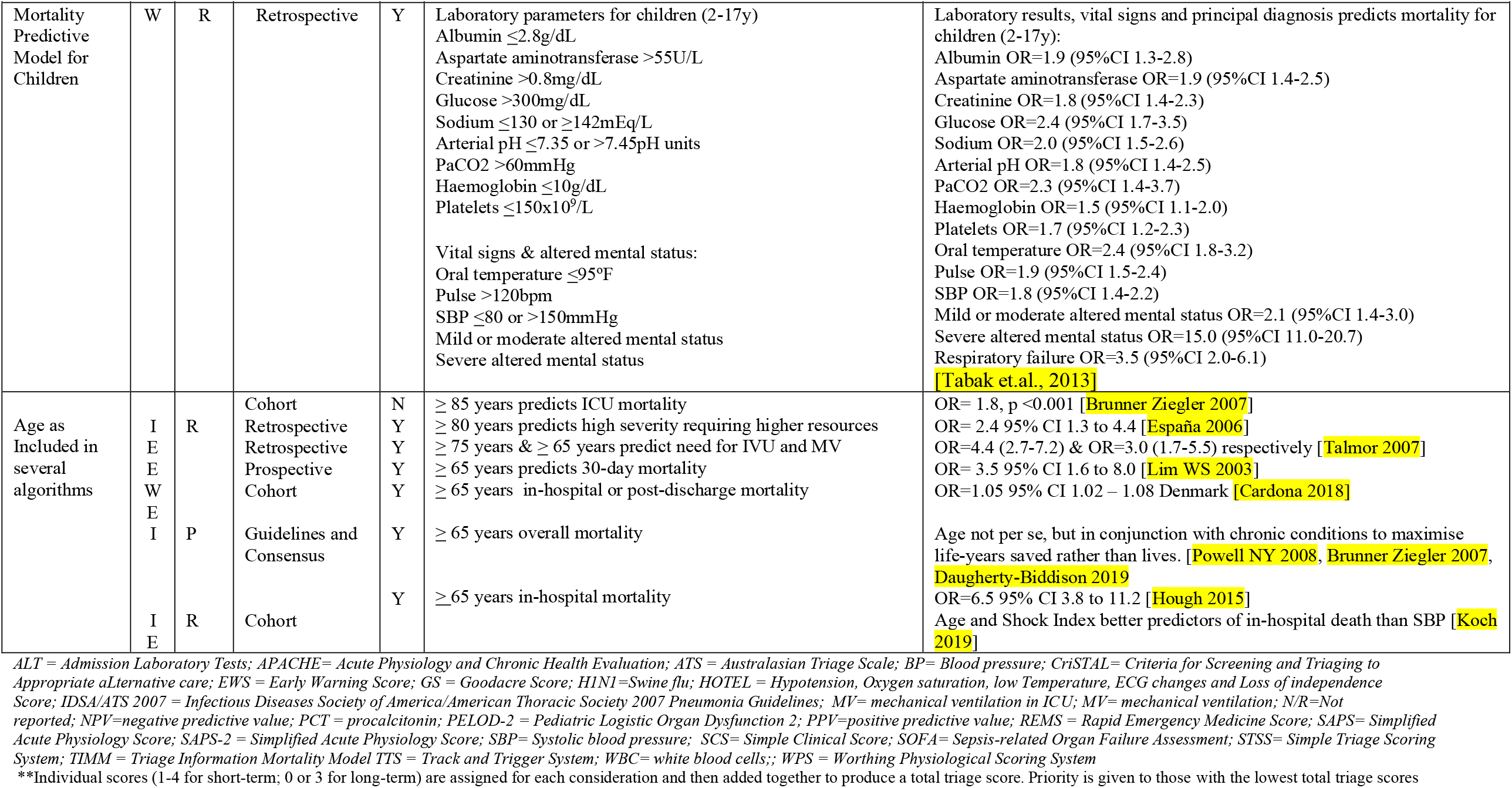
**Validation status, and clinical parameters predicting poor outcome for potentially guidance on ICU admission or discharge derived from <u>routine care</u> studies in ICU, ED or wards(N= 46 studies)**

## References

1. Tanne JH; Hayasaki E; Zastrow M; Pulla P; Smith P; Garcia RA. Covid-19: how doctors and healthcare systems are tackling coronavirus worldwide. BMJ 2020, 368, m1090.

2. Maves, R.C.; Downar, J.; Dichter, J.R.; Hick, J.L.; Devereaux, A.; Geiling, J.A.; Kissoon, N.; Hupert, N.; Niven, A.S.; King, M.A.; et al. Triage of Scarce Critical Care Resources in COVID-19 An Implementation Guide for Regional Allocation: An Expert Panel Report of the Task Force for Mass Critical Care and the American College of Chest Physicians. Chest 2020, 158, 212–225, doi:10.1016/j.chest.2020.03.063.

3. Aziz, S.; Arabi, Y.M.; Alhazzani, W.; Evans, L.; Citerio, G.; Fischkoff, K.; Salluh, J.; Meyfroidt, G.; Alshamsi, F.; Oczkowski, S.; et al. Managing ICU surge during the COVID-19 crisis: rapid guidelines. Intensive Care Med 2020, 10.1007/s00134-020-06092-5, 1–23, doi: 10.1007/s00134-020-06092-5.

4. Immovilli, P.; Morelli, N.; Antonucci, E.; Radaelli, G.; Barbera, M.; Guidetti, D. COVID-19 mortality and ICU admission: the Italian experience. Crit Care 2020, 24, 228–228, doi:10.1186/s13054-020-02957-9.

5. Mahase, E. Covid-19: most patients require mechanical ventilation in first 24 hours of critical care. BMJ 2020, 368, m1201, doi:10.1136/bmj.m1201.

6. Auld, S.; Caridi-Scheible, M.; Blum, J.M.; Robichaux, C.J.; Kraft, C.S.; Jacob, J.T.; Jabaley, C.S.; Carpenter, D.; Kaplow, R.; Hernandez, A.C.; et al. ICU and ventilator mortality among critically ill adults with COVID-19. Crit Care Med 2020, Online First, doi:10.1097/CCM.0000000000004457.

7. Grasselli, G.; Zangrillo, A.; Zanella, A.; Antonelli, M.; Cabrini, L.; Castelli, A.; Cereda, D.; Coluccello, A.; Foti, G.; Fumagalli, R.; et al. Baseline Characteristics and Outcomes of 1591 Patients Infected With SARS-CoV-2 Admitted to ICUs of the Lombardy Region, Italy. Jama 2020, 323, 1574–1581, doi:10.1001/jama.2020.5394.

8. Cummings, M.J.; Baldwin, M.R.; Abrams, D.; Jacobson, S.D.; Meyer, B.J.; Balough, E.M.; Aaron, J.G.; Claassen, J.; Rabbani, L.E.; Hastie, J.; et al. Epidemiology, clinical course, and outcomes of critically ill adults with COVID-19 in New York City: a prospective cohort study. Lancet (London, England) 2020, 395, 1763–1770, doi:10.1016/S0140-6736(20)31189-2.

9. Ioannidis, J.P.A.; Axfors, C.; Contopoulos-Ioannidis, D.G. Population-level COVID-19 mortality risk for non-elderly individuals overall and for non-elderly individuals without underlying diseases in pandemic epicenters. medRxiv 2020, 10.1101/2020.04.05.20054361, 2020.2004.2005.20054361, doi:10.1101/2020.04.05.20054361.

10. Toltzis, P.; Soto-Campos, G.; Shelton, C.; Kuhn, E.M.; Hahn, R.; Kanter, R.K.; Wetzel, R.C. Evidence-Based Pediatric Outcome Predictors to Guide the Allocation of Critical Care Resources in a Mass Casualty Event. Pediatric critical care medicine: a journal of the Society of Critical Care Medicine and the World Federation of Pediatric Intensive and Critical Care Societies 2015, 16, e207–216, doi:10.1097/pcc.0000000000000481.

11. Christian, M.D.; Hawryluck, L.; Wax, R.S.; Cook, T.; Lazar, N.M.; Herridge, M.S.; Muller, M.P.; Gowans, D.R.; Fortier, W.; Burkle, F.M. Development of a triage protocol for critical care during an influenza pandemic. Cmaj 2006, 175, 1377–1381.

12. Antommaria, A.H.; Sweney, J.; Poss, W.B. Critical appraisal of: triaging pediatric critical care resources during a pandemic: ethical and medical considerations. Pediatric critical care medicine: a journal of the Society of Critical Care Medicine and the World Federation of Pediatric Intensive and Critical Care Societies 2010, 11, 396–400, doi:10.1097/PCC.0b013e3181dac698.

13. Cardona M; Anstey M; Lewis ET; Shanmugam S; Hillman K. Appropriateness of intensive care treatments near the end of life during the COVID-19 pandemic. Breathe 2020, Sept, In Press, doi:https://doi.org/10.1183/20734735.0062-2020.

14. LeDuc, J.W.; Barry, M.A. SARS, the First Pandemic of the 21st Century. Emerg Infect Dis 2004, 10, e26–e26, doi:10.3201/eid1011.040797_02.

15. Devereaux, A.V.; Dichter, J.R.; Christian, M.D.; Dubler, N.N.; Sandrock, C.E.; Hick, J.L.; Powell, T.; Geiling, J.A.; Amundson, D.E.; Baudendistel, T.E.; et al. Definitive care for the critically ill during a disaster: a framework for allocation of scarce resources in mass critical care: from a Task Force for Mass Critical Care summit meeting, January 26-27, 2007, Chicago, IL. Chest 2008, 133, 51s–66s, doi:10.1378/chest.07-2693.

16. New York State Task Force in Life and The Law. Ventilator allocation guidelines; 2015.

17. Powell, T.; Christ, K.C.; Birkhead, G.S. Allocation of ventilators in a public health disaster. Disaster med 2008, 2, 20–26, doi:https://dx.doi.org/10.1097/DMP.0b013e3181620794.

18. White, D.B.; Katz, M.H.; Luce, J.M.; Lo, B. Who should receive life support during a public health emergency? Using ethical principles to improve allocation decisions. Ann Intern Med 2009, 150, 132–138, doi:10.7326/0003-4819-150-2-200901200-00011.

19. Wilkens, E.P.; Klein, G.M. Mechanical ventilation in disaster situations: a new paradigm using the AGILITIES Score System. Am J Disaster Med 2010, 5, 369–384.

20. Chuang, E.; Cuartas, P.A.; Powell, T.; Gong, M.N. “We’re Not Ready, But I Don’t Think You’re Ever Ready.” Clinician Perspectives on Implementation of Crisis Standards of Care. AJOB Empirical Bioethics 2020, 11, 148–159, doi:10.1080/23294515.2020.1759731.

21. Adeniji, K.A.; Cusack, R. The Simple Triage Scoring System (STSS) successfully predicts mortality and critical care resource utilization in H1N1 pandemic flu: a retrospective analysis. Crit Care 2011, 15, R39, doi:10.1186/cc10001.

22. Grissom, C.K.; Brown, S.M.; Kuttler, K.G.; Boltax, J.P.; Jones, J.; Jephson, A.R.; Orme, J.F., Jr. A modified sequential organ failure assessment score for critical care triage. Disaster Med Public Health Prep 2010, 4, 277–284, doi:10.1001/dmp.2010.40.

23. Guest, T.; Tantam, G.; Donlin, N.; Tantam, K.; McMillan, H.; Tillyard, A. An observational cohort study of triage for critical care provision during pandemic influenza: ‘clipboard physicians’ or ‘evidenced based medicine’? Anaesthesia 2009, 64, 1199–1206, doi:10.1111/j.1365-2044.2009.06084.x.

24. Khan, Z.; Hulme, J.; Sherwood, N. An assessment of the validity of SOFA score based triage in H1N1 critically ill patients during an influenza pandemic. Anaesthesia 2009, 64, 1283–1288, doi:10.1111/j.1365-2044.2009.06135.x.

25. Liang, W.; Liang, H.; Ou, L.; Chen, B.; Chen, A.; Li, C.; Li, Y.; Guan, W.; Sang, L.; Lu, J.; et al. Development and Validation of a Clinical Risk Score to Predict the Occurrence of Critical Illness in Hospitalized Patients With COVID-19. JAMA Intern Med 2020, 180, 1–9, doi:10.1001/jamainternmed.2020.2033.

26. Semple, M.G.; Myles, P.R.; Nicholson, K.G.; Lim, W.S.; Read, R.C.; Taylor, B.L.; Brett, S.J.; Openshaw, P.J.; Enstone, J.E.; McMenamin, J.; et al. An evaluation of community assessment tools (CATs) in predicting use of clinical interventions and severe outcomes during the A(H1N1)pdm09 pandemic. PloS one 2013, 8, e75384, doi:10.1371/journal.pone.0075384.

27. Talmor, D.; Jones, A.E.; Rubinson, L.; Howell, M.D.; Shapiro, N.I. Simple triage scoring system predicting death and the need for critical care resources for use during epidemics. Crit Care Med 2007, 35, 1251–1256, doi:10.1097/01.Ccm.0000262385.95721.Cc.

28. Kim, K.M.; Cinti, S.; Gay, S.; Goold, S.; Barnosky, A.; Lozon, M. Triage of mechanical ventilation for pediatric patients during a pandemic. Disaster med 2012, 6, 131–137, doi:https://dx.doi.org/10.1001/dmp.2012.19.

29. Daugherty-Biddison, E.L.; Faden, R.; Gwon, H.S.; Mareiniss, D.P.; Regenberg, A.C.; Schoch-Spana, M.; Schwartz, J.; Toner, E.S. Too Many Patients…A Framework to Guide Statewide Allocation of Scarce Mechanical Ventilation During Disasters. Chest 2019, 155, 848–854, doi:https://dx.doi.org/10.1016/j.chest.2018.09.025.

30. Dólera-Moreno, C.; Palazón-Bru, A.; Colomina-Climent, F.; Gil-Guillén, V.F. Construction and internal validation of a new mortality risk score for patients admitted to the intensive care unit. Int J Clin Pract 2016, 70, 916–922, doi:10.1111/ijcp.12851.

31. Granholm, A.; Perner, A.; Krag, M.; Hjortrup, P.B.; Haase, N.; Holst, L.B.; Marker, S.; Collet, M.O.; Jensen, A.K.G.; Møller, M.H. Development and internal validation of the Simplified Mortality Score for the Intensive Care Unit (SMS-ICU). Acta Anaesthesiol Scand 2018, 62, 336–346, doi:10.1111/aas.13048.

32. Gupta, P.; Rettiganti, M.; Gossett, J.M.; Daufeldt, J.; Rice, T.B.; Wetzel, R.C. Development and Validation of an Empiric Tool to Predict Favorable Neurologic Outcomes Among PICU Patients. Crit Care Med 2018, 46, 108–115, doi:10.1097/ccm.0000000000002753.

33. Hajjar, L.A.; Nakamura, R.E.; de Almeida, J.P.; Fukushima, J.T.; Hoff, P.M.; Vincent, J.L.; Auler, J.O., Jr.; Galas, F.R. Lactate and base deficit are predictors of mortality in critically ill patients with cancer. Clinics (Sao Paulo) 2011, 66, 2037–2042, doi:10.1590/s1807-59322011001200007.

34. Slater, A.; Shann, F.; Pearson, G. PIM2: a revised version of the Paediatric Index of Mortality. Intensive Care Med 2003, 29, 278–285, doi:10.1007/s00134-002-1601-2.

35. Aegerter, P.; Boumendil, A.; Retbi, A.; Minvielle, E.; Dervaux, B.; Guidet, B. SAPS II revisited. Intensive Care Med 2005, 31, 416–423, doi:10.1007/s00134-005-2557-9.

36. Brunner-Ziegler, S.; Heinze, G.; Ryffel, M.; Kompatscher, M.; Slany, J.; Valentin, A. “Oldest old” patients in intensive care: prognosis and therapeutic activity. Wien Klin Wochenschr 2007, 119, 14–19, doi:10.1007/s00508-007-0771-x.

37. Charles, P.G.; Wolfe, R.; Whitby, M.; Fine, M.J.; Fuller, A.J.; Stirling, R.; Wright, A.A.; Ramirez, J.A.; Christiansen, K.J.; Waterer, G.W.; et al. SMART-COP: a tool for predicting the need for intensive respiratory or vasopressor support in community-acquired pneumonia. Clinical infectious diseases: an official publication of the Infectious Diseases Society of America 2008, 47, 375–384, doi:10.1086/589754.

38. Higgins, T.L.; Teres, D.; Nathanson, B. Outcome prediction in critical care: the Mortality Probability Models. Curr Opin Crit Care 2008, 14, 498–505, doi:10.1097/MCC.0b013e3283101643.

39. Hough, C.L.; Caldwell, E.S.; Cox, C.E.; Douglas, I.S.; Kahn, J.M.; White, D.B.; Seeley, E.J.; Bangdiwala, S.I.; Rubenfeld, G.D.; Angus, D.C.; et al. Development and Validation of a Mortality Prediction Model for Patients Receiving 14 Days of Mechanical Ventilation. Crit Care Med 2015, 43, 2339–2345, doi:10.1097/ccm.0000000000001205.

40. Na, H.J.; Jeong, E.S.; Kim, I.; Kim, W.Y.; Lee, K. Clinical Application of the Quick Sepsis-Related Organ Failure Assessment Score at Intensive Care Unit Admission in Patients with Bacteremia: A Single-Center Experience of Korea. Korean J Crit Care Med 2017, 32, 247–255, doi:10.4266/kjccm.2017.00241.

41. Nielsen, A.B.; Thorsen-Meyer, H.C.; Belling, K.; Nielsen, A.P.; Thomas, C.E.; Chmura, P.J.; Lademann, M.; Moseley, P.L.; Heimann, M.; Dybdahl, L.; et al. Survival prediction in intensive-care units based on aggregation of long-term disease history and acute physiology: a retrospective study of the Danish National Patient Registry and electronic patient records. The Lancet Digital Health 2019, 1, e78–e89, doi:10.1016/S2589-7500(19)30024-X.

42. Pannu, S.R.; Moreno Franco, P.; Li, G.; Malinchoc, M.; Wilson, G.; Gajic, O. Development and validation of severe hypoxemia associated risk prediction model in 1,000 mechanically ventilated patients*. Crit Care Med 2015, 43, 308–317, doi:10.1097/ccm.0000000000000671.

43. Godfrey, G.; Pilcher, D.; Hilton, A.; Bailey, M.; Hodgson, C.L.; Bellomo, R. Treatment limitations at admission to intensive care units in Australia and New Zealand: prevalence, outcomes, and resource use*. Crit Care Med 2012, 40, 2082–2089, doi:10.1097/CCM.0b013e31824ea045.

44. Rosjo, H.; Stridsberg, M.; Ottesen, A.H.; Christensen, G.; Petilla, V.; Linko, R.; Karlsson, S.; Varpula, T.; Ruokonen, E.; Omland, T. The novel cardiovascular biomarker secretoneurin predicts mortality and shock in critical ill patients with infections. European Heart Journal 2015, 36, 1053, doi:10.1093/eurheartj/ehv401.

45. Schuetz, P.; Maurer, P.; Punjabi, V.; Desai, A.; Amin, D.N.; Gluck, E. Procalcitonin decrease over 72 hours in US critical care units predicts fatal outcome in sepsis patients. Crit Care 2013, 17, R115, doi:10.1186/cc12787.

46. Sprung, C.L.; Baras, M.; Iapichino, G.; Kesecioglu, J.; Lippert, A.; Hargreaves, C.; Pezzi, A.; Pirracchio, R.; Edbrooke, D.L.; Pesenti, A.; et al. The Eldicus prospective, observational study of triage decision making in European intensive care units: part I--European Intensive Care Admission Triage Scores. Crit Care Med 2012, 40, 125–131, doi:10.1097/CCM.0b013e31822e5692.

47. Hartley, T.M.; Lane, N.D.; Steer, J.; Elliott, M.W.; Sovani, M.; Curtis, H.J.; Fuller, E.R.; Murphy, P.B.; Hart, N.; Shrikrishna, D.; et al. Predicting outcome from exacerbations of COPD requiring assisted ventilation: Results from the NIV outcome (NIVO) study. Thorax 2019, 74, A38–A39, doi:10.1136/thorax-2019-BTSabstracts2019.63.

48. Kristensen, M.; Iversen, A.K.S.; Gerds, T.A.; Østervig, R.; Linnet, J.D.; Barfod, C.; Lange, K.H.W.; Sölétormos, G.; Forberg, J.L.; Eugen-Olsen, J.; et al. Routine blood tests are associated with short term mortality and can improve emergency department triage: a cohort study of >12,000 patients. Scand J Trauma Resusc Emerg Med 2017, 25, 115, doi:10.1186/s13049-017-0458-x.

49. Mark, K.; George, N.; Rasheed, H.; Meurer, D.; Allen, B.; Elie-Turenne, M.C.; Hou, P.; Seethala, R. QSOFA criteria predicts clinical outcomes of hospitalized emergency department pneumonia patients. Crit Care Med 2016, 44, 407, doi:10.1097/01.ccm.0000510001.55136.6c.

50. O’Sullivan, E.; Callely, E.; O’Riordan, D.; Bennett, K.; Silke, B. Predicting outcomes in emergency medical admissions - role of laboratory data and co-morbidity. Acute Med 2012, 11, 59–65.

51. Singer, M.; Deutschman, C.S.; Seymour, C.W.; Shankar-Hari, M.; Annane, D.; Bauer, M.; Bellomo, R.; Bernard, G.R.; Chiche, J.D.; Coopersmith, C.M.; et al. The Third International Consensus Definitions for Sepsis and Septic Shock (Sepsis-3). Jama 2016, 315, 801–810, doi:10.1001/jama.2016.0287.

52. Soong, J.T.Y.; Poots, A.; Rolph, G.; Bell, D. Frailty syndromes coded within secondary user service(SUS) data predict inpatient mortality and long length of stay. Acute Medicine 2017, 16, 131–132.

53. Brabrand, M.; Folkestad, L.; Clausen, N.G.; Knudsen, T.; Hallas, J. Risk scoring systems for adults admitted to the emergency department: a systematic review. Scand J Trauma Resusc Emerg Med 2010, 18, 8, doi:10.1186/1757-7241-18-8.

54. España, P.P.; Capelastegui, A.; Gorordo, I.; Esteban, C.; Oribe, M.; Ortega, M.; Bilbao, A.; Quintana, J.M. Development and validation of a clinical prediction rule for severe community-acquired pneumonia. American journal of respiratory and critical care medicine 2006, 174, 1249–1256, doi:10.1164/rccm.200602-177OC.

55. Jalbout, N.A.; Hamade, B.; Balhara, K.S.; Hsieh, Y.H.; Bayram, J.D. Shock index as a predictor of hospital admission and inpatient mortality in a united states national database of emergency departments. Journal of Emergency Medicine 2017, 53, 433, doi:10.1016/j.jemermed.2017.08.040.

56. Koch, E.; Lovett, S.; Nghiem, T.; Riggs, R.A.; Rech, M.A. Shock index in the emergency department: utility and limitations. Open access emergency medicine: OAEM 2019, 11, 179–199, doi:10.2147/oaem.S178358.

57. Teubner, D.J.; Considine, J.; Hakendorf, P.; Kim, S.; Bersten, A.D. Model to predict inpatient mortality from information gathered at presentation to an emergency department: The Triage Information Mortality Model (TIMM). Emerg Med Australas 2015, 27, 300–306, doi:10.1111/1742-6723.12425.

58. Brown, S.M.; Jones, J.P.; Aronsky, D.; Jones, B.E.; Lanspa, M.J.; Dean, N.C. Relationships among initial hospital triage, disease progression and mortality in community-acquired pneumonia. Respirology 2012, 17, 1207–1213, doi:10.1111/j.1440-1843.2012.02225.x.

59. Grossmann, F.F.; Zumbrunn, T.; Frauchiger, A.; Delport, K.; Bingisser, R.; Nickel, C.H. At risk of undertriage? Testing the performance and accuracy of the emergency severity index in older emergency department patients. Ann Emerg Med 2012, 60, 317-325.e313, doi:10.1016/j.annemergmed.2011.12.013.

60. Kellett, J.; Deane, B. The Simple Clinical Score predicts mortality for 30 days after admission to an acute medical unit. Qjm 2006, 99, 771–781, doi:10.1093/qjmed/hcl112.

61. Subbe, C.P.; Jishi, F.; Hibbs, R.A. The simple clinical score: a tool for benchmarking of emergency admissions in acute internal medicine. Clinical medicine (London, England) 2010, 10, 352–357, doi:10.7861/clinmedicine.10-4-352.

62. Yu, S.; Leung, S.; Heo, M.; Soto, G.J.; Shah, R.T.; Gunda, S.; Gong, M.N. Comparison of risk prediction scoring systems for ward patients: a retrospective nested case-control study. Crit Care 2014, 18, R132, doi:10.1186/cc13947.

63. Allegretti, A.S.; Steele, D.J.; David-Kasdan, J.A.; Bajwa, E.; Niles, J.L.; Bhan, I. Continuous renal replacement therapy outcomes in acute kidney injury and end-stage renal disease: a cohort study. Crit Care 2013, 17, R109, doi:10.1186/cc12780.

64. Bethea, A.; Seidler, D.; Coleman, C.; Johnson, K.; Davis, E.; Thompson, S. Prehospital identification of elevated lactic acid levels and sepsis-related outcomes (lasr study). Crit Care Med 2018, 46, 687, doi:10.1097/01.ccm.0000529407.05908.9b.

65. Cardona, M.; Lewis, E.T.; Kristensen, M.R.; Skjøt-Arkil, H.; Ekmann, A.A.; Nygaard, H.H.; Jensen, J.J.; Jensen, R.O.; Pedersen, J.L.; Turner, R.M.; et al. Predictive validity of the CriSTAL tool for short-term mortality in older people presenting at Emergency Departments: a prospective study. Eur Geriatr Med 2018, 9, 891–901, doi:10.1007/s41999-018-0123-6.

66. Haas, L.; Eckart, A.; Haubitz, S.; Mueller, B.; Schuetz, P.; Segerer, S. Estimated glomerular filtration rate predicts 30-day mortality in medical emergency departments: Results of a prospective multi-national observational study. PloS one 2020, 15, e0230998–e0230998, doi:10.1371/journal.pone.0230998.

67. Hutchison, C.A.; Crowe, A.V.; Stevens, P.E.; Harrison, D.A.; Lipkin, G.W. Case mix, outcome and activity for patients admitted to intensive care units requiring chronic renal dialysis: a secondary analysis of the ICNARC Case Mix Programme Database. Crit Care 2007, 11, R50, doi:10.1186/cc5785.

68. Wallis, S.J.; Wall, J.; Biram, R.W.; Romero-Ortuno, R. Association of the clinical frailty scale with hospital outcomes. Qjm 2015, 108, 943–949, doi:10.1093/qjmed/hcv066.

69. Lim, W.S.; van der Eerden, M.M.; Laing, R.; Boersma, W.G.; Karalus, N.; Town, G.I.; Lewis, S.A.; Macfarlane, J.T. Defining community acquired pneumonia severity on presentation to hospital: an international derivation and validation study. Thorax 2003, 58, 377, doi:10.1136/thorax.58.5.377.

70. Marwick, C.A.; Guthrie, B.; Pringle, J.E.; McLeod, S.R.; Evans, J.M.; Davey, P.G. Identifying which septic patients have increased mortality risk using severity scores: a cohort study. BMC Anesthesiol 2014, 14, 1, doi:10.1186/1471-2253-14-1.

71. Stræde, M.; Brabrand, M. External validation of the simple clinical score and the HOTEL score, two scores for predicting short-term mortality after admission to an acute medical unit. PloS one 2014, 9, e105695, doi:10.1371/journal.pone.0105695.

72. Tabak, Y.P.; Sun, X.; Hyde, L.; Yaitanes, A.; Derby, K.; Johannes, R.S. Using enriched observational data to develop and validate age-specific mortality risk adjustment models for hospitalized pediatric patients. Med Care 2013, 51, 437–445, doi:10.1097/MLR.0b013e318287d57d.

73. van Walraven C; McAlister FA; Bakal JA; Hawken S; Donzé J. External validation of the Hospital-patient One-year Mortality Risk (HOMR) model for predicting death within 1 year after hospital admission. Cmaj 2015, 187, 725–733.

74. Heyland, D.K.; Davidson, J.; Skrobik, Y.; des Ordons, A.R.; Van Scoy, L.J.; Day, A.G.; Vandall-Walker, V.; Marshall, A.P. Improving partnerships with family members of ICU patients: study protocol for a randomized controlled trial. Trials 2018, 19, 3, doi:10.1186/s13063-017-2379-4.

75. Einav, S.; Hick, J.L.; Hanfling, D.; Erstad, B.L.; Toner, E.S.; Branson, R.D.; Kanter, R.K.; Kissoon, N.; Dichter, J.R.; Devereaux, A.V.; et al. Surge capacity logistics: care of the critically ill and injured during pandemics and disasters: CHEST consensus statement. Chest 2014, 146, e17S–43S, doi:https://dx.doi.org/10.1378/chest.14-0734.

76. Murray, S.A.; Kendall, M.; Boyd, K.; Sheikh, A. Illness trajectories and palliative care. BMJ (Clinical research ed.) 2005, 330, 1007–1011, doi:10.1136/bmj.330.7498.1007.

77. Heyland, D.K.; Stelfox, H.T.; Garland, A.; Cook, D.; Dodek, P.; Kutsogiannis, J.; Jiang, X.; Turgeon, A.F.; Day, A.G.; on behalf of the Canadian Critical Care Trials, G., et al. Predicting Performance Status 1 Year After Critical Illness in Patients 80 Years or Older: Development of a Multivariable Clinical Prediction Model. Crit Care Med 2016, 44.

78. Adeniji, K.; Cusack, R.; Golder, K. The simple triage scoring system (STSS) successfully predicts mortality and critical care resource utilisation in H1N1 pandemic flu. Intensive Care Medicine 2010, 36, S286, doi:10.1007/s00134-010-2000-8.

79. World Health Organization. Clinical management of COVID-19. Interim guidance; WHO/2019-nCoV/clinical/2020.5; 2020; p 62.

80. Australia and New Zealand Intensive Care Society. [web report] Guiding principles for complex decision making during Pandemic COVID-19. Version 2.0. Availabe online: https://www.anzics.com.au/wp-content/uploads/2020/04/ANZI_3367_Guidelines_V2.pdf (accessed on June).

81. Winsor, S.; Bensimon, C.M.; Sibbald, R.; Anstey, K.; Chidwick, P.; Coughlin, K.; Cox, P.; Fowler, R.; Godkin, D.; Greenberg, R.A.; et al. Identifying prioritization criteria to supplement critical care triage protocols for the allocation of ventilators during a pandemic influenza. Healthc Q 2014, 17, 44–51.

82. Minne, L.; Abu-Hanna, A.; de Jonge, E. Evaluation of SOFA-based models for predicting mortality in the ICU: A systematic review. Crit Care 2008, 12, R161, doi:10.1186/cc7160.

83. Alam, A.; Gupta, S. Lactate Measurements and Their Association With Mortality in Pediatric Severe Sepsis in India: Evidence That 6-Hour Level Performs Best. J Intensive Care Med 2020, 10.1177/0885066620903231, 885066620903231, doi:10.1177/0885066620903231.

84. Bozcuk, H.; Koyuncu, E.; Yildiz, M.; Samur, M.; Ozdogan, M.; Artaç, M.; Coban, E.; Savas, B. A simple and accurate prediction model to estimate the intrahospital mortality risk of hospitalised cancer patients. Int J Clin Pract 2004, 58, 1014–1019, doi:10.1111/j.1742-1241.2004.00169.x.

85. Rothstein, M.A. Currents in contemporary ethics. Should health care providers get treatment priority in an influenza pandemic? J Law Med Ethics 2010, 38, 412–419, doi:10.1111/j.1748-720X.2010.00499.x.

86. Daugherty, E.L.; Rubinson, L. Preparing your intensive care unit to respond in crisis: considerations for critical care clinicians. Crit Care Med 2011, 39, 2534–2539, doi:https://dx.doi.org/10.1097/CCM.0b013e3182326440.

87. Patel JJ; Heyland DK. Unmasking the triumphs, tragedies and opportunities of the COVID-19 pandemic. ICU Management and Practice 2020, 3.

88. Van Scoy, L.J.; Chiarolanzio, P.J.; Kim, C.; Heyland, D.K. Development and initial evaluation of an online decision support tool for families of patients with critical illness: A multicenter pilot study. J Crit Care 2017, 39, 18–24, doi:10.1016/j.jcrc.2016.12.022.

89. Bion J; Dennis A. ICU admission and discharge criteria. In Oxford Textbook of critical care, 2nd ed., Oxford University Press: 2016; 10.1093/med/9780199600830.003.0020.

90. Mareiniss, D.P.; Levy, F.; Regan, L. ICU triage: the potential legal liability of withdrawing ICU care during a catastrophic event. Am J Disaster Med 2011, 6, 329–338.

91. Hick JL; Hanfling D; Wynia MK; Pavia AT. Duty to Plan: Health Care, Crisis Standards of Care, and Novel Coronavirus SARS-CoV-2. NAM Perspectives. Discussion paper. National Academy of Medicine 2020, https://doi.org/10.31478/202003b, doi:https://doi.org/10.31478/202003b.

92. Fadul, N.; Elsayem, A.F.; Bruera, E. Integration of palliative care into COVID-19 pandemic planning. BMJ Supportive & Palliative Care 2020, 10.1136/bmjspcare-2020-002364, bmjspcare-2020-002364, doi:10.1136/bmjspcare-2020-002364.

93. New York State Task Force on Life & the Law. Ventilator allocation guidelines. New York State Department of Health: New York, USA, 2015; p 266 pp.

94. Biddison, L.D.; Berkowitz, K.A.; Courtney, B.; De Jong, C.M.; Devereaux, A.V.; Kissoon, N.; Roxland, B.E.; Sprung, C.L.; Dichter, J.R.; Christian, M.D.; et al. Ethical considerations: care of the critically ill and injured during pandemics and disasters: CHEST consensus statement. Chest 2014, 146, e145S–155S, doi:https://dx.doi.org/10.1378/chest.14-0742.

95. Clayton, J.M.; Hancock, K.M.; Butow, P.N.; Tattersall, M.H.; Currow, D.C.; Adler, J.; Aranda, S.; Auret, K.; Boyle, F.; Britton, A.; et al. Clinical practice guidelines for communicating prognosis and end-of-life issues with adults in the advanced stages of a life-limiting illness, and their caregivers. The Medical journal of Australia 2007, 186, S77–s105.

96. Mercadante, S.; Gregoretti, C.; Cortegiani, A. Palliative care in intensive care units: why, where, what, who, when, how. BMC Anesthesiology 2018, 18, 106, doi:10.1186/s12871-018-0574-9.

97. Braganza MA; Glossop A; Vora VA. Treatment withdrawal and end-of-life care in the intensive care unit. BJA Education 2017, 17, 396–400.

98. Silva, D.S.; Gibson, J.L.; Robertson, A.; Bensimon, C.M.; Sahni, S.; Maunula, L.; Smith, M.J. Priority setting of ICU resources in an influenza pandemic: a qualitative study of the Canadian public’s perspectives. BMC public health 2012, 12, 241, doi:10.1186/1471-2458-12-241.

99. Silva, D.S.; Nie, J.X.; Rossiter, K.; Sahni, S.; Upshur, R.E. Contextualizing ethics: ventilators, H1N1 and marginalized populations. Healthcare quarterly (Toronto, Ont.) 2010, 13, 32–36, doi:10.12927/hcq.2013.21613.

100. Australasian College of Emergency Medicine. Clinical Guidelines for the management of COVID-19 in AUstralasian Emergency Depratments. Availabe online: https://acem.org.au/getmedia/78105c4b-5195-43f6-9c91-25dda5604eaf/Clinical-Guidelines (accessed on v3.0).

101. Australia and New Zealand Intensive Care Society. COVID-19 Guidelines version 2. Availabe online: https://www.anzics.com.au/wp-content/uploads/2020/04/ANZI_3367_Guidelines_V2.pdf (accessed on

102. Sprung, C.L.; Zimmerman, J.L.; Christian, M.D.; Joynt, G.M.; Hick, J.L.; Taylor, B.; Richards, G.A.; Sandrock, C.; Cohen, R.; Adini, B.; et al. Recommendations for intensive care unit and hospital preparations for an influenza epidemic or mass disaster: summary report of the European Society of Intensive Care Medicine’s Task Force for intensive care unit triage during an influenza epidemic or mass disaster. Intensive Care Med 2010, 36, 428–443, doi:https://dx.doi.org/10.1007/s00134-010-1759-y.

103. Challen, K.; Bentley, A.; Bright, J.; Walter, D. Clinical review: mass casualty triage--pandemic influenza and critical care. Crit Care 2007, 11, 212.

104. Haniffa, R.; Mukaka, M.; Munasinghe, S.B.; De Silva, A.P.; Jayasinghe, K.S.A.; Beane, A.; de Keizer, N.; Dondorp, A.M. Simplified prognostic model for critically ill patients in resource limited settings in South Asia. Crit Care 2017, 21, 250, doi:10.1186/s13054-017-1843-6.

